# Genomic Epidemiology of *Salmonella and Campylobacter* in Poultry Production: Quantifying the Contribution of Primary Breeders

**DOI:** 10.1101/2025.11.11.25340000

**Authors:** David J Lipman

**Affiliations:** Department of Pathology, Stanford University School of Medicine, Stanford, CA, USA; Department of Biomedical Engineering, Johns Hopkins University, Baltimore, Maryland; Center for Computational Biology, Johns Hopkins University, Baltimore, Maryland

## Abstract

The US broiler production system processes over 9.3 billion chickens annually through a highly integrated pyramid structure where two primary breeding companies supply genetic stock to approximately 40 major integrators operating nationwide. To provide a quantitative, system-wide estimate of contamination origins, I analyzed whole genome sequences from *Salmonella* and *Campylobacter* isolates collected from over 800 processing facilities as part of the USDA’s Food Safety Inspection Service (FSIS) verification sampling (2019-2024). Single-linkage clustering identified isolates sharing common origins (≤2, 4, or 8 SNPs genome-wide), which were categorized by facility, company, and geographical distributions to infer contamination sources. Among the isolates analyzed, geographically dispersed multi-company clusters implying a primary breeder origin accounted for 78% of *Campylobacter*, 77% of non-Enteritidis *Salmonella*, and 96% of *Salmonella* Enteritidis isolates. Geographic spread analysis revealed that Enteritidis isolates matched a random distribution model consistent with contamination originating from the highest levels of the breeding pyramid. *Campylobacter* showed regional clustering implying sources at lower levels of the breeding pyramid. Cluster persistence exceeded multiple production cycles (median >4 years for *Campylobacter*, >4.5 years for 75% of Enteritidis isolates), indicating stable contamination reservoirs upstream of processing. These results demonstrate that the primary breeders are a major source of broiler contamination and suggest that upstream interventions targeting breeding stock, in particular for Enteritidis, may represent an efficient strategy for further reducing clinical cases of foodborne illness.

## Introduction

Poultry is a major source of foodborne illness in the United States accounting for approximately 24-33% of the cases of *Salmonella* infection (CDC 2025; Rose et al. 2025) and 50-70% of the cases of *Campylobacter* infection (Cody et al. 2019; Wilson et al. 2008; Pascoe et al. 2024). The Federal Meat Inspection Act of 1906 (“Federal Meat Inspection Act,” n.d.) provided the mandate for the United States Department of Agriculture (USDA) to inspect animal carcasses at processing complexes (“U.S.C. Title 21 - FOOD AND DRUGS,” n.d.) and that process has evolved as our understanding of food safety and the associated methodologies have progressed (Johnston 2020). Since 2013, whole genome sequencing (WGS) of pathogens has been used by the Centers for Disease Control (CDC), Food and Drug Administration (FDA), and the USDA to detect outbreaks of foodborne illness and identify their contamination source (Jackson et al. 2016). More recently, the Food Safety and Inspection Service (FSIS) of the United States Department of Agriculture (USDA) has also been using WGS in their raw poultry sampling program (“Laboratory Sampling Data,” n.d.) as part of their overall efforts to establish performance standards and improve risk analysis.

The value of WGS in food safety is based on the premise that when the genome sequences of a set of isolates differ by only a small number of mutations (e.g., a threshold of 4 or fewer single nucleotide polymorphisms, or SNPs for the entire genome sequence), there is a strong probability that these isolates share a common contamination source (Brown et al. 2019; Jagadeesan et al. 2019; Ronholm et al. 2016; E. L. Stevens et al. 2022; Pightling et al. 2018). This can be applied to a set of clinical isolates to detect a foodborne outbreak and, when including environmental isolates, to help trace the likely contamination source. A recent analytical approach based on clusters of isolates grouped by a threshold SNP distance has been used to gain new insights on sporadic food poisoning (Lipman et al. 2024). This same approach can be used to analyze the *Salmonella* and *Campylobacter* isolates from the FSIS poultry carcass sampling program to improve our understanding of the origin within the production system for the pathogens that contaminate the broilers.

The broiler production system in the United States is often compared to a pyramid (see **Figure 1**) where a relatively tiny number of chickens is expanded in multiple generations to generate the over 9.5 billion chickens slaughtered in 2024 (United States Department of Agriculture, n.d.). Though there are hundreds of smaller poultry production companies, by 2013, 95% of the broiler chickens in the United States were produced by approximately 29,500 farmers under contract with 40 vertically integrated companies (Iowa State University, {Animal and Plant Health Inspection Service], and USDA, n.d.). For a typical processing complex, the poultry integrators generally own the processing plants as well as the associated feed mills, hatcheries, and trucking. These integrators contract out for the associated breeder farms and growout houses clustered within the overall complex near the processing plant, primarily using exclusive agreements (Iowa State University, {Animal and Plant Health Inspection Service], and USDA, n.d.; MacDonald 2008). And just two primary breeder companies provide Parent hatchlings to the breeder farms on complexes owned or controlled by these major integrators (Iowa State University, {Animal and Plant Health Inspection Service], and USDA, n.d.; Akilian 2020).

**Figure 1.**
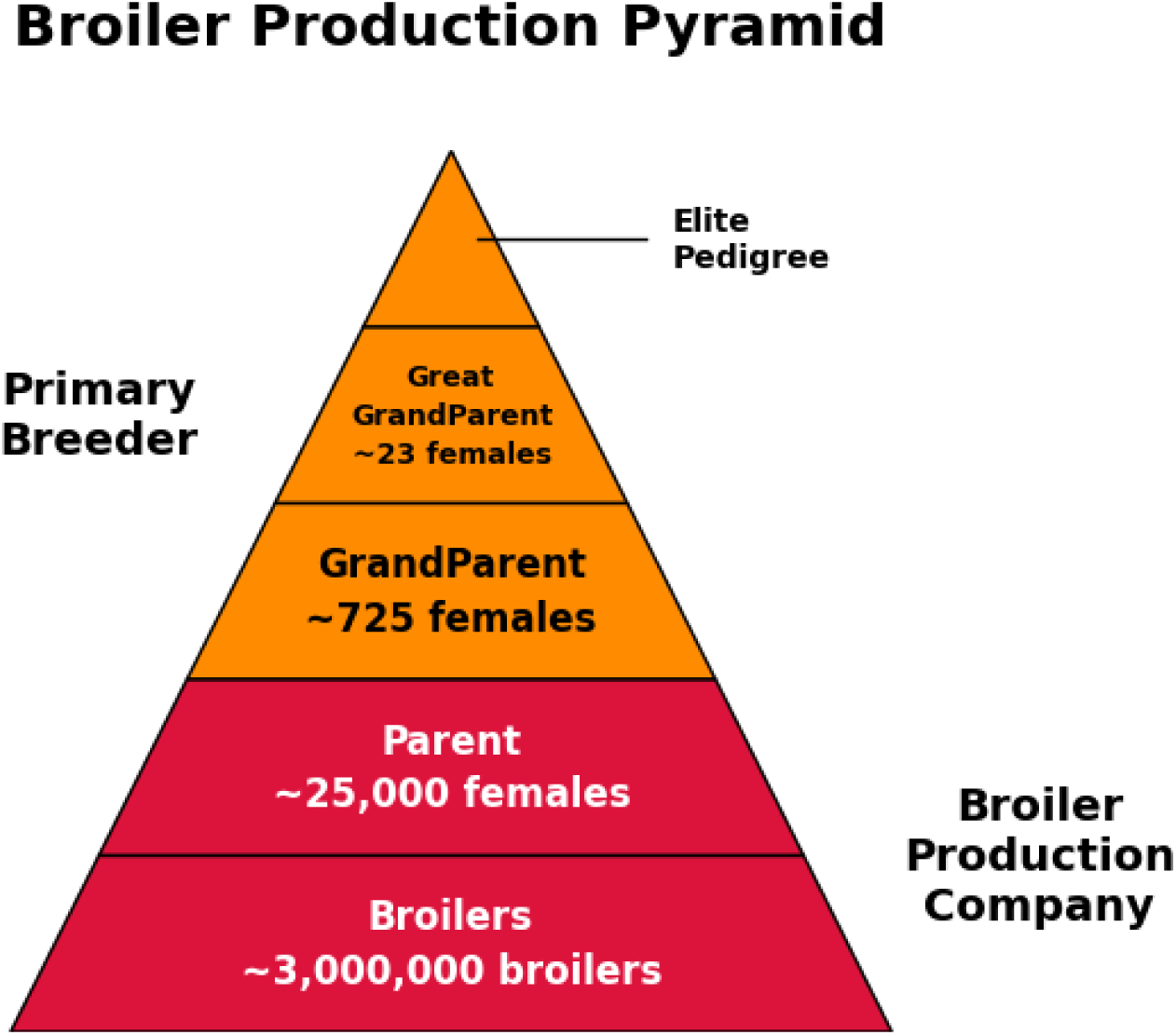
Broiler Production Pyramid. In orange are the generations of the breeding pyramid managed by the primary breeder companies and in red are those of the companies that manage broiler production and processing within the processing complexes (Pollock 1999; Van Eenennaam et al. 2014).

There have been extensive analyses of the sources of Salmonella and Campylobacter contamination in the various phases of broiler production. In one study of three processing complexes in the United States, the prevalence of Salmonella at the live receiving end of the process was > 85% (Chavez-Velado, Vargas, and Sanchez-Plata 2024). A systematic review of the literature on Campylobacter in poultry estimated that live birds had a predicted prevalence of 41% (Wang et al. 2023a). The bacterial populations can change significantly at later phases of processing (Wang et al. 2023a, 2023b; Golden and Mishra 2020; Chavez-Velado, Vargas, and Sanchez-Plata 2024; Williams et al. 2025). Our analysis focuses on samples from the post-chill stage of broiler production, as this is where the FSIS collects samples for verification testing of *Salmonella* and *Campylobacter*, following all processing interventions (“FSIS Compliance Guideline: Modernization of Poultry Slaughter Inspection Microbiological Sampling of Raw Poultry” 2015).

Metadata from each FSIS sample includes the collection date, the processing facility identifier and its state, as well as the name of the company that owns the complex. Thus, in examining a cluster of isolates with nearly identical genomes, we can note whether all came from the same complex, different complexes but owned by the same processing company, different companies but same state, and different companies and from different states. What inferences can we make based on these relationships?

One source of the contamination found in the FSIS verification testing samples would be within the various components of the broiler processing complexes themselves. For the health of their flocks and their customers, the companies that own and control these complexes have strong incentives to minimize the presence of pathogens within their processing complexes. The National Poultry Improvement Plan (NPIP), a cooperative industry, state, and federal program, was initially established in 1935 to reduce the incidence of Pullorum Disease which caused high mortality rates among flocks and a number of biosecurity standards were developed and implemented within these processing complexes (“Animal Health,” n.d.; Iowa State University, {Animal and Plant Health Inspection Service], and USDA, n.d.). To minimize horizontal transmission of pathogens, the locations of these processing complexes are chosen to be geographically isolated from other broiler processing complexes. A number of steps are taken to sanitize vehicles, limit personnel access, minimize contamination from feed, and implement active pest control (USDA, The Center for Food Security and Public Health, Iowa State University, n.d.). Additional guidelines have been widely implemented such as the Perimeter Buffer Area (PBA) separating poultry houses from areas not involved in production, the Line of Separation (LOS) which is a functional line reducing exposure of the chickens within a poultry house (National Poultry Improvement Plan, n.d.), and the “All in/All out” approach for extensive sanitation between the processing of flocks (Usda, n.d.; USDA, Iowa State University Center for Food Security and Public Health 2013).

Contamination in FSIS samples can also originate from Parent chicks transferred from primary breeder companies (*upstream* phase) to Parent farms within broiler processing complexes (*downstream* phase), as illustrated in **Figure 2**. Each of the two primary breeder companies supplies multiple processing complexes, which can be owned by different companies and located in multiple states. Specifically, a single primary breeder GrandParent farm might supply Parent chicks to several different processor companies’ Parent farms. This means that contamination at this stage of the primary breeder system could lead to the transmission of pathogen strains to a number of complexes across several nearby states. Furthermore, due to the structure of the breeding pyramid, contaminating strains that emerge at earlier stages of the primary breeder system, such as the Great GrandParent stage, have the potential to spread to multiple GrandParent farms of that primary breeder. In turn, these strains could then be disseminated to a large number of processing complexes owned by different companies and located throughout the country (see **Figure 2**). And as noted above, contamination introduced earlier in the overall process could be eliminated at later stages.

**Figure 2.**
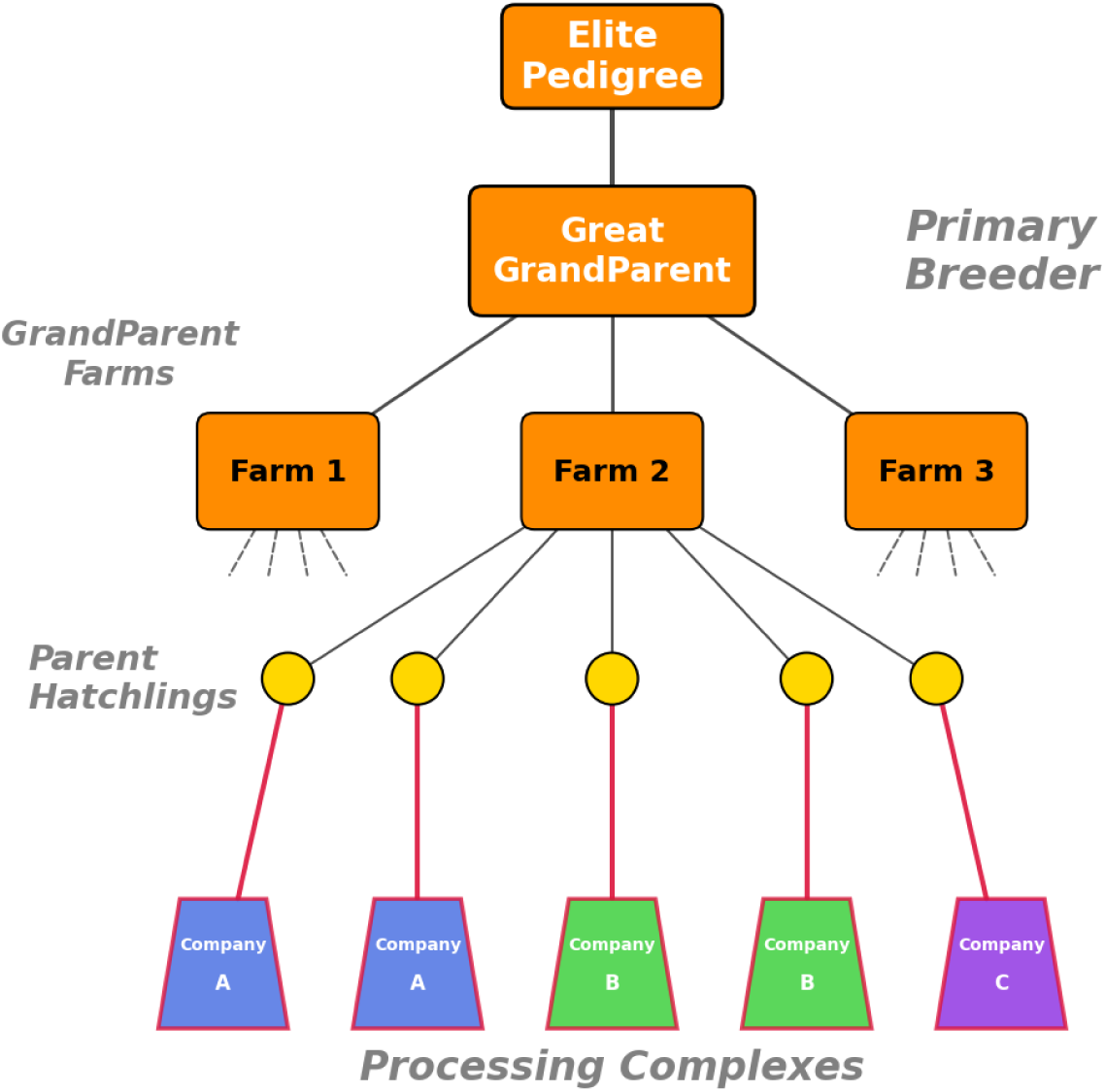
The Tree Structure of the Breeding Pyramid. In orange are the levels controlled by the primary breeder and outlined in red, those components of the poultry processors. Each GrandParent Farm serves different geographical regions and provides Parent Hatchlings to processing complexes managed by multiple companies and located in multiple states.

A pathogen strain introduced in the upstream phase could be found in isolates from numerous chickens across multiple processing complexes, owned by different companies and located in multiple states. Conversely, a strain originating in the downstream phase, because of the extensive measures associated with the NPIP and related initiatives, would more likely be found in relatively fewer chickens and within a single processing complex. However, if a single company operates multiple complexes within the same region, then there may be some opportunities to introduce the same strain to these complexes. Even when attempts are made to isolate processing complexes, the proximity of multiple complexes—which may be owned by different companies—means that wildlife, such as birds or insects, could still introduce the same strain of pathogen to these nearby facilities.

Since 2019, isolates from the positive FSIS verification testing samples have undergone whole genome sequencing. I have generated clusters formed using stringent SNP thresholds (e.g. 4 SNPs across the entire genome) from the FSIS sequenced *Salmonella* and *Campylobacter* isolates to determine isolates that likely share a common source of contamination. I then characterized these clusters in terms of the processing complexes, companies, and states associated with isolates in the cluster. From these relationships and our knowledge of the structure of the integrated broiler production system, we can make inferences about isolates that are likely to be from contamination introduced upstream at the primary breeder stage or downstream within the processor complex.

Consider two contrasting models at opposite ends of the spectrum:

1. **Internal Contamination:** If all contamination originates within the processing complex, each isolate in a cluster would be from the same processor.
2. **Early-Stage Contamination:** If all contamination stems from an early stage of the breeding pyramid within the primary breeder, each cluster would effectively be a random sample drawn from the total set of facilities along with their associated companies and geographic locations.

As noted above, even if most of the contamination were from internal sources, nearby complexes could share clusters from the same environmental source or an integrator company could inadvertently spread contamination among its own complexes. Likewise, if the contamination originates, e.g. within a GrandParent farm of the primary breeder rather than a Great GrandParent farm, then the facilities represented within a cluster may be more geographically restricted. Furthermore, if some facilities are more successful than others at eliminating contamination introduced by the primary breeder in their broilers or if some pathogen clusters show more horizontal spread within some facilities than others, then the dispersion pattern for these clusters will differ from the random model described in #2 above.

For the isolates that I can categorize, i.e. those in clusters greater than size 1, the analysis indicates that the upstream stage, i.e., the primary breeder, is the source of approximately 78% of *Campylobacter* isolates, more than 96% of *Salmonella* Enteritidis isolates, and over 77% of the *Salmonella* (excluding Enteritidis) isolates. Furthermore, most Enteritidis isolates originate from the earliest stages of the primary breeder pyramid.

## Results

### Basic properties of dataset

**Figure 3** shows the number of monthly broiler samples at processing facilities around the US (blue bars, axis on left side), the number of samples that yielded Campylobacter isolates (green line, y axis on right side), and the number yielding Salmonella isolates (red line, y axis on right side). Note that in October 2023, FSIS decreased the fraction of samples being tested for Campylobacter (USDA/FSIS, n.d.) such that the actual fraction of positives has stayed relatively constant.

**Figure 3.**
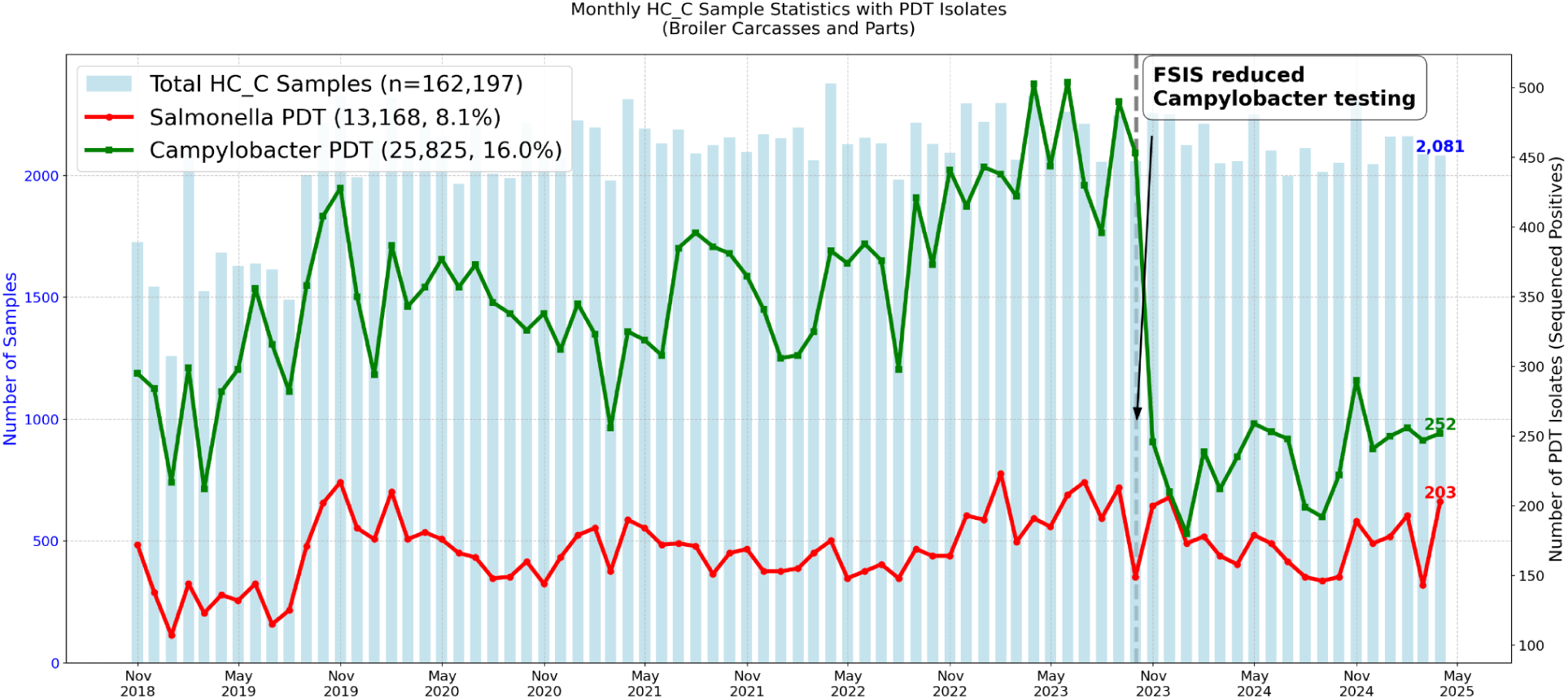
Monthly FSIS samples and associated *Campylobacter* and *Salmonella* genomes. The blue bars represent the number of FSIS samples per month (y axis on left). In green are the number of monthly *Campylobacter* genomes from the positive samples (y axis on right) and in red are the number of monthly *Salmonella* genomes.

**Figure 4** provides a summary of a portion of the metadata and shows that most samples are being taken from a relatively small number of companies. These companies have multiple processing complexes in multiple states.

**Figure 4.**
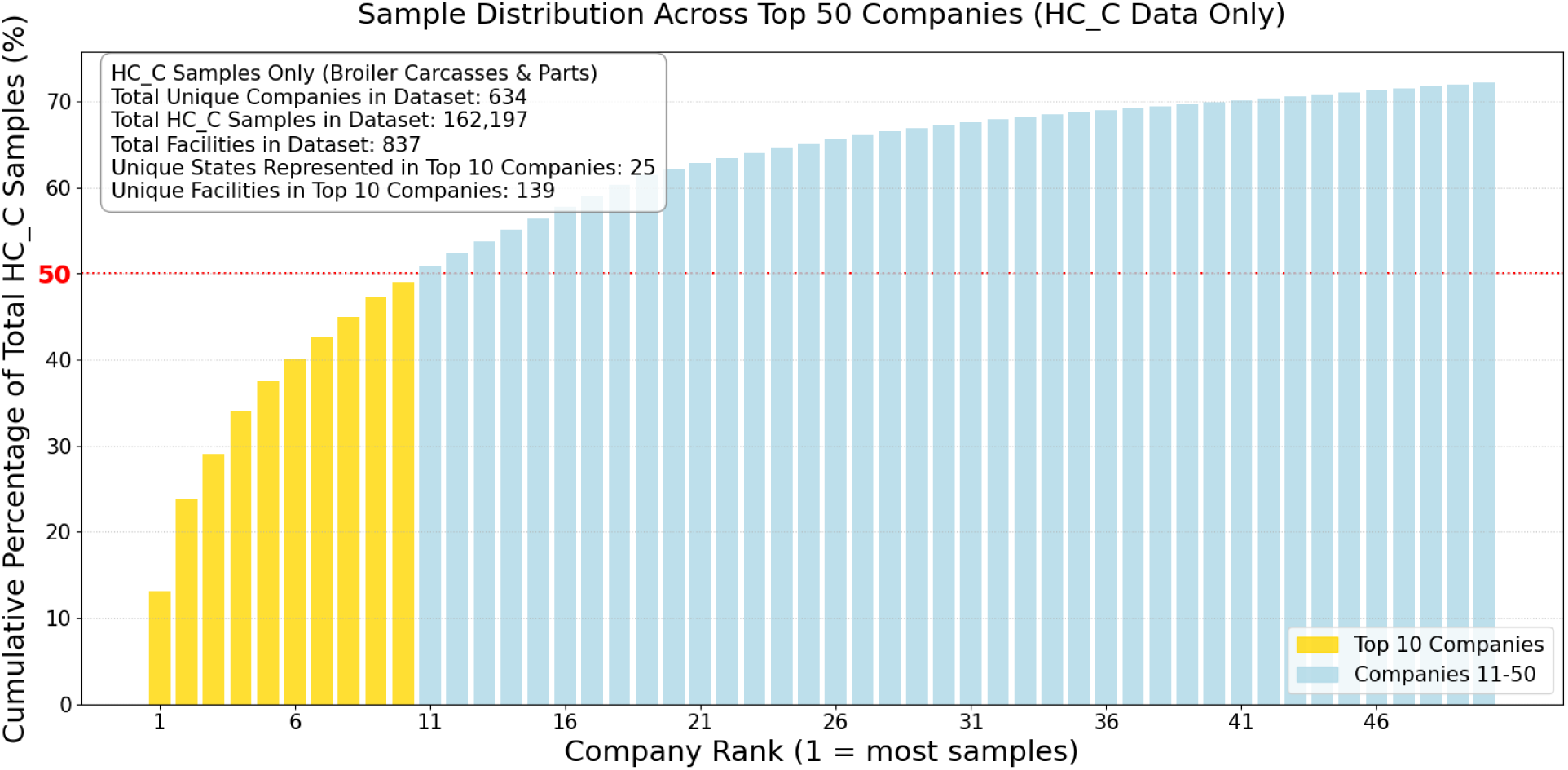
Sample Distribution across Top 50 Companies. Vertical bars show the cumulative percentage of samples from the companies with the most samples. The top 10 companies are in yellow.

There are 64 *Salmonella* serovars among the sequenced isolates and the six most common serovars are:

1. Infantis, 29.6%,
2. Kentucky, 27.3%,
3. Enteritidis, 19.4%,
4. Typhimurium, 9.2%,
5. Schwarzengrund, 4.9%

For *Campylobacter*, 47% are *Campylobacter jejuni* and 53% are *Campylobacter coli*.

Clusters of poultry isolates were generated as described in Lipman et al. (Lipman et al. 2024) and are shown in **Figure 5**. Briefly, I use a threshold SNP distance for pairs of genomes that is low enough to infer a high likelihood that the associated pathogen isolates are derived from the same source of contamination, e.g., four SNPs out of ∼2 to 5 million aligned bases. I continue to add cases to the cluster as long as they are within this threshold distance from at least one case already in the cluster, i.e., single linkage clustering. The results presented here are based on a threshold of four SNPs; however using higher or lower thresholds did not qualitatively change the results (**Figure S1**).

**Figure 5.**
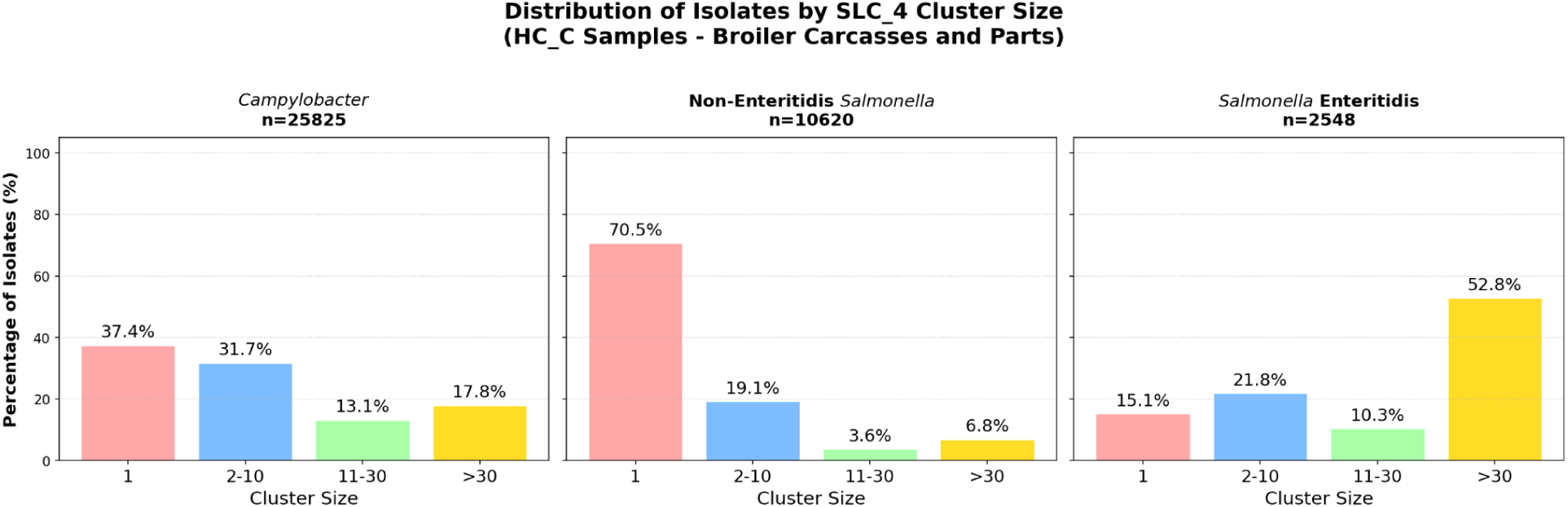
Distribution of isolates by cluster size for *Campylobacter*, non-Enteritidis *Salmonella*, and Enteritidis. Vertical bars represent the proportion of isolates in the cluster size ranges indicated on the x axis for each pathogen group.

*Salmonella* Enteritidis isolates are in much larger clusters than Campylobacter or non-Enteritidis *Salmonella*. For example, while only about 31% of *Campylobacter* and ∼11% of non-Enteritidis *Salmonella* isolates are in clusters larger than size 10, ∼63% of the Enteritidis isolates are in clusters larger than size 10.

**Figure 6** shows the locations of 81 complexes owned by 52 companies across the United States where samples yielded isolates from a single *Salmonella* cluster. In this figure, the geographic resolution is at the level of state so placement within the states is arbitrary. For the five companies with the largest number of complexes within the cluster I indicate their complexes with a colored marker. For the rest of the complexes, I only indicate the number of complexes in each state. The geographic distribution of the processing complexes for isolates within the same cluster will be analyzed quantitatively below.

**Figure 6.**
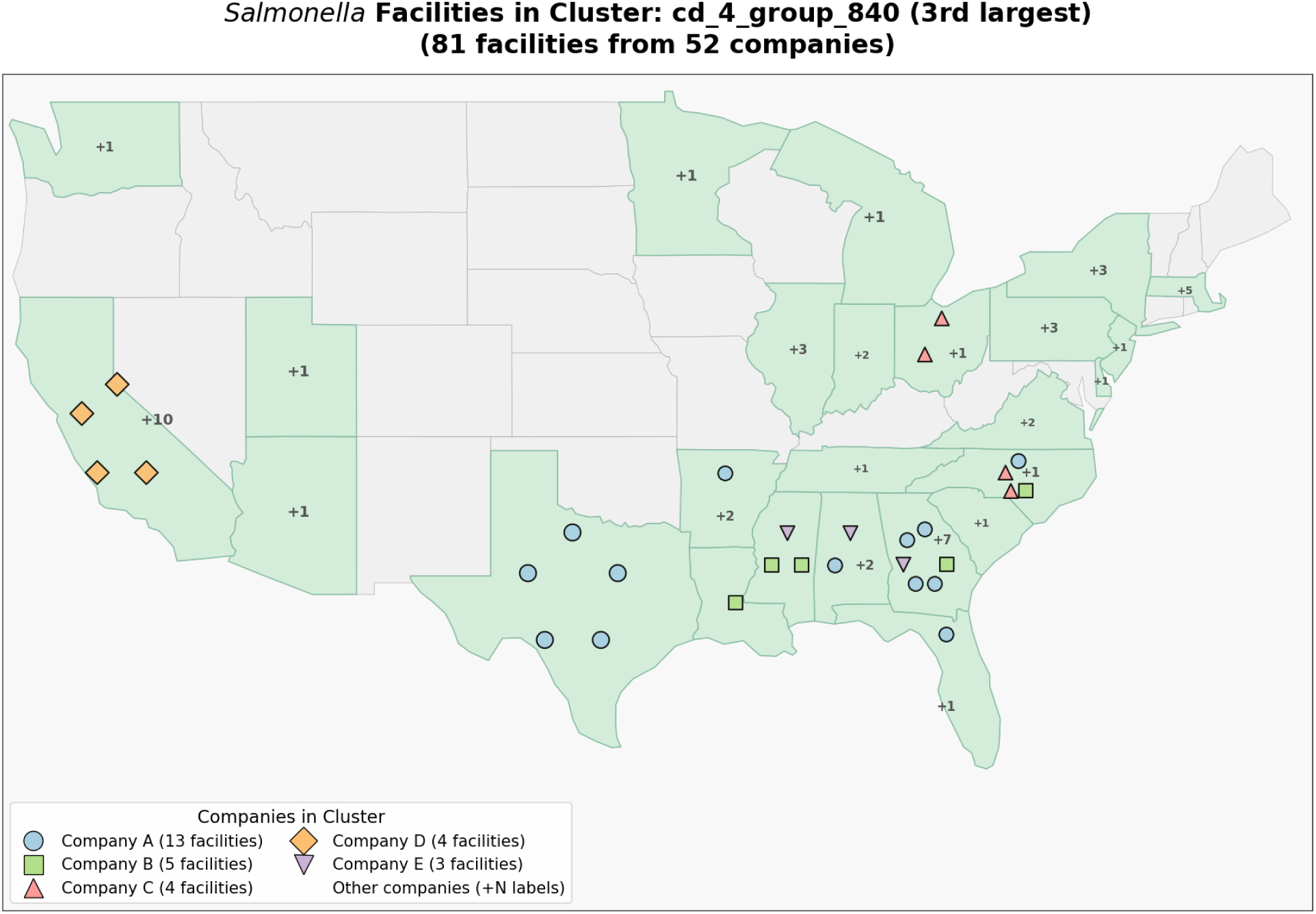
Geographic spread for a single large *Salmonella* cluster. The locations for the isolates from a single Salmonella cluster are shown. Positions within the state borders are arbitrary and spaced for visibility. Colored markers are provided for the companies with the largest number of facilities in the cluster. The digits indicate the number of additional facilities within each state.

**Figure 7** illustrates the sample collection timeline for a *Salmonella* cluster and a comparable timeline for a *Campylobacter* cluster. Each dot signifies a positive sample collected on a specific date, where the isolate belonged to that particular cluster. Different complexes are differentiated by color, and if a single complex has multiple samples collected on various dates, these dots are interconnected by lines. The line’s height denotes the cumulative sample count from that facility at a given time point. For both clusters, isolates were found across different complexes, companies, and states over several years, and at the same complex for extended durations. Since the first and last isolates in these two clusters were collected near the sampling interval’s boundaries, some isolates may be unaccounted for since they occurred before or after our sampling interval. As can be seen in **Figure 7**, a cluster can persist for multiple years in the same facility and at multiple different facilities during the same time period. Cluster persistence will be analyzed quantitatively below.

**Figure 7.**
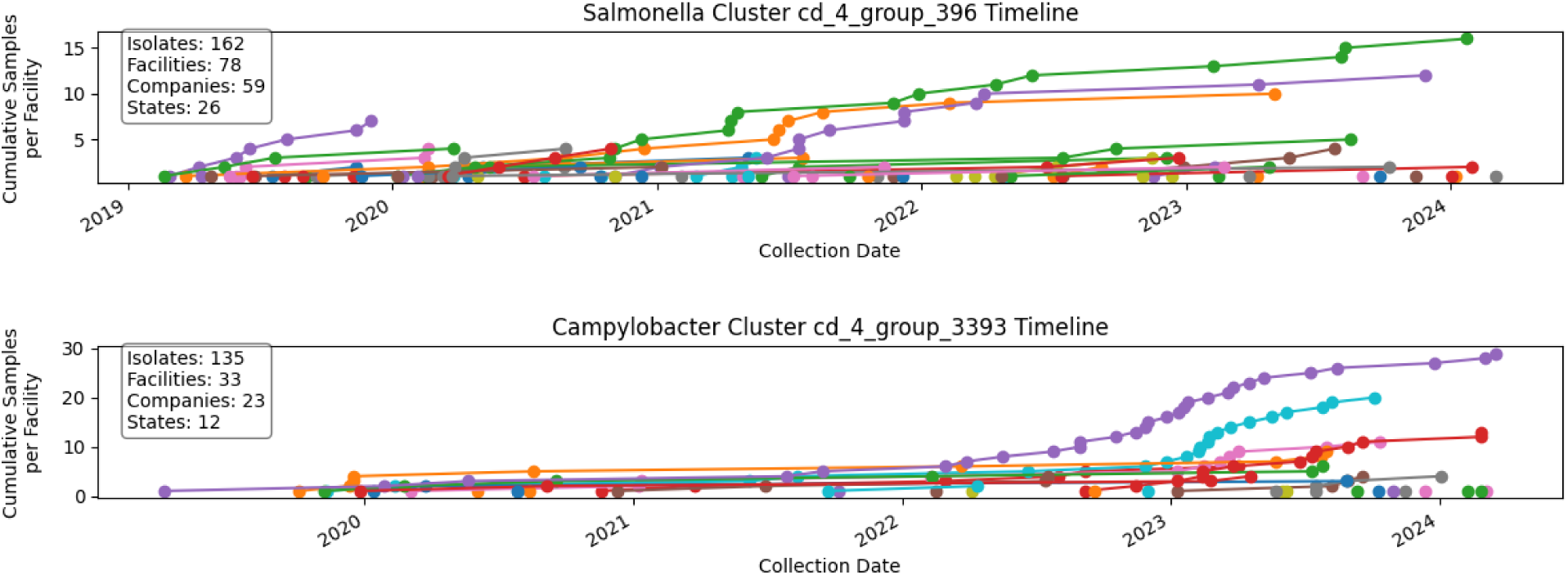
Timeline of samples from a single cluster. The top panel displays a timeline of samples within a single *Salmonella* cluster. Circles of the same color indicate samples originating from a single facility, with colored lines connecting multiple sample dates from that facility. The cumulative number of samples per facility is represented by the height of these connected circles. The bottom panel illustrates a similar sampling timeline, but for a single *Campylobacter* cluster.

### Cluster categories

Clusters of two or more isolates can be classified based on the metadata of their constituent samples. These spread categories are as follows:

- **Single Complex**: All isolates originate from the same processing complex.
- **Multiple Complexes, Single Company**: Isolates come from multiple complexes but only one company.
- **Multiple Companies:** Isolates come from complexes that are owned by different companies.

**Figure 8** shows the percentages of isolates in each of the three categories for *Campylobacter*, Enteritidis, and non-Enteritidis *Salmonella*. Most of the isolates of all three pathogen groups are in multi-company clusters though a significant fraction of Campylobacter and non-Enteritidis *Salmonella* isolates are in single complex clusters. And very few isolates are in the multi-complex, single company clusters. Using a 2 SNP threshold for clustering slightly decreases the fraction of isolates in multi-company clusters and an 8 SNP threshold slightly increases this fraction at the expense of the single facility clusters (see **Figure S2**).

**Figure 8.**
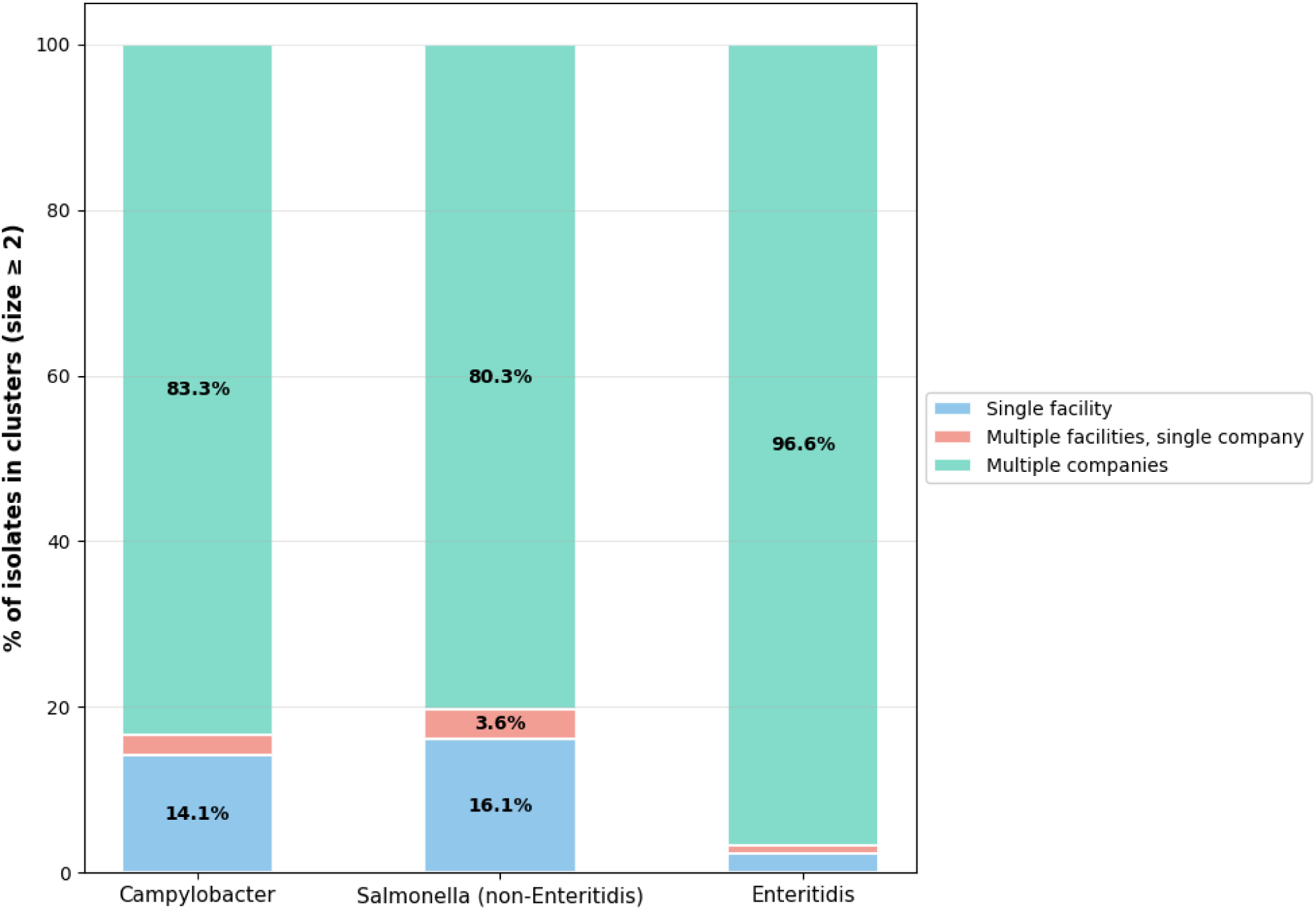
Proportion of isolates in each spread category for *Campylobacter*, Enteritidis, non-Enteritidis *Salmonella*. Stacked bar plots showing the proportion of isolates in single facility, multi-facility and single company, and multi-company clusters for *Campylobacter*, Enteritidis, and non_Enteritidis *Salmonella*.

One can characterize the geographic spread of isolates within each multi-company cluster by using the latitude and longitude of the associated processing complexes. To assess the likelihood of a local environmental source for the shared contamination within a cluster, I will determine the maximum pairwise distance among the isolates for that cluster. The mean pairwise distance will be used to characterize the cluster’s overall geographic spread. The first panel in **Figure 9** shows the fraction of isolates in clusters with increasing maximum pairwise distance for the three pathogen groups. For all three pathogen groups, most isolates are in clusters with large maximum pairwise distances. A maximum pairwise distance of 50 miles, for instance, encompasses a minimum circular area of nearly 2,000 square miles, yet most isolates are found in clusters with significantly greater maximum pairwise distances. Moreover, the smooth decline shared by the three pathogen curves offers no clear boundary that would signal a transition away from local contamination sources. Note that virtually identical results are obtained computing the mean pairwise distance within clusters **(Figure S4).**

**Figure 9.**
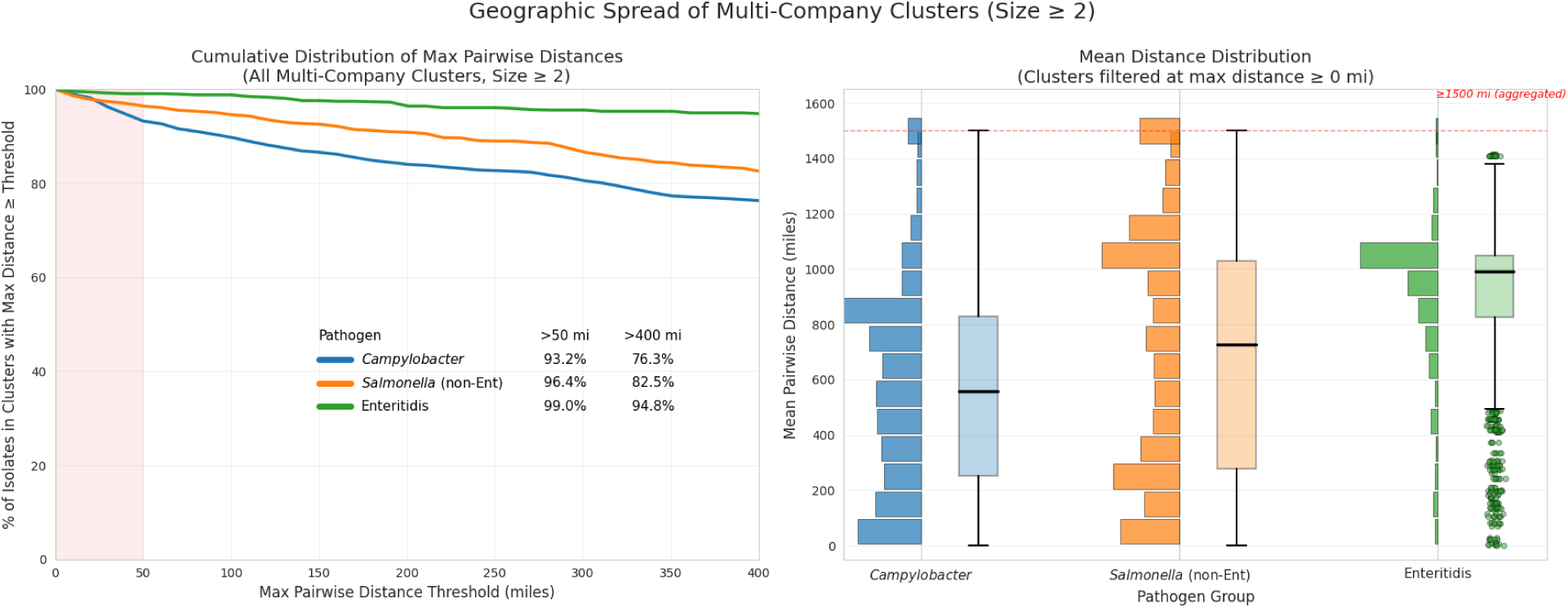
Distribution of Pairwise Distances within Multi-Company Clusters. The left panel shows cumulative distribution curves of the proportion of isolates in clusters with increasing maximum pairwise distance between processing complexes. The right panel shows histograms and box plots for the distribution of the proportion of isolates in clusters with different mean pairwise distances. The y axis was truncated at 1500 miles and counts for isolates in clusters with mean distance > 1500 are aggregated in the last bin of the histograms.

The second panel in **Figure 9** illustrates the distribution of the proportion of isolates found in clusters with varying mean pairwise distances. Enteritidis isolates exhibit the greatest geographical spread followed by non-Enteritidis *Salmonella* and *Campylobacter*. Notably, the histograms for both non-Enteritidis *Salmonella* and *Campylobacter* display a bimodal distribution, indicating a significantly higher proportion of regionally concentrated isolates compared to Enteritidis. The geographical dispersion within clusters is highly robust, as evidenced by the nearly identical results in **Figure S3** regardless of whether 2, 4, or 8 SNP thresholds were used for clustering.

To analyze the dispersion of isolates in multi-company clusters, I examined all isolate pairs within each cluster. This allowed us to determine which pairs were from different complexes or different companies, in addition to calculating the mean pairwise distance for these isolate pairs (**Figure 10**). To determine a spread ratio, I compare our measures to those from a random model. This random model represents the expectation if all processing complexes sampled from a single, shared contamination source. It is constructed by shuffling all isolates among the complexes while preserving the original cluster memberships and the links between complexes and their respective companies. For all comparisons, Enteritidis isolates are approximately as dispersed as the random model with non-Enteritidis *Salmonella* and *Campylobacter* showing less dispersion than the random model. Consistent with **Figure 9**, we can see that non-Enteritidis *Salmonella* and in particular, *Campylobacter* show reduced geographic dispersion while there is relatively less reduction of dispersion among facilities and companies. Clustering using either a 2 SNP or 8 SNP threshold yields nearly identical results (**Figure S5)**. This further demonstrates the robustness of these findings regarding dispersion within clusters.

**Figure 10.**
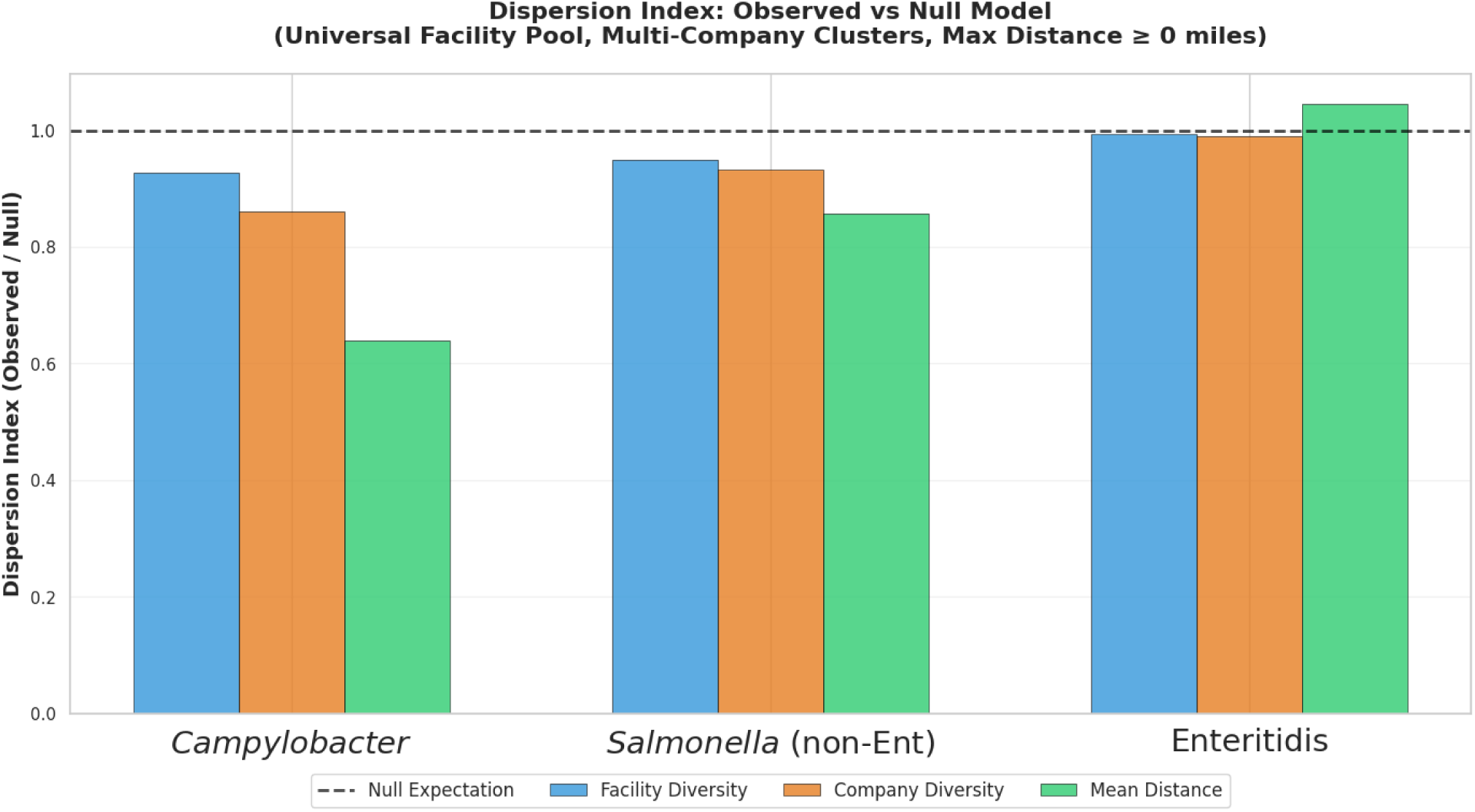
Spread Ratio of Observed versus Random Pair Counts for Multi-Company Clusters. The vertical bars represent the ratios of observed counts versus random pair counts (50 iterations): all isolate pairs within each cluster are examined to determine whether they are from different complexes (blue bar) or different companies (orange), and the mean pairwise distance for the cluster is computed. These counts are compared to the same counts for the random reference model.

**Figure 11** displays box plots showing the log scale distribution of cluster sizes of the isolates in single company versus multi-company clusters. Notably, isolates in multi-company clusters have much larger cluster sizes. The Enteritidis multi-company cluster sizes are particularly large which is consistent with what has been reported for Enteritidis isolates from clinical cases (Lipman et al. 2024). Results using thresholds of 2 SNPs or 8 SNPs for clustering are qualitatively similar (**Figure S6**) with the distributions for multi-company isolates shifted somewhat to smaller cluster sizes with 2 SNP thresholds and to larger sizes with 8 SNP thresholds.

**Figure 11.**
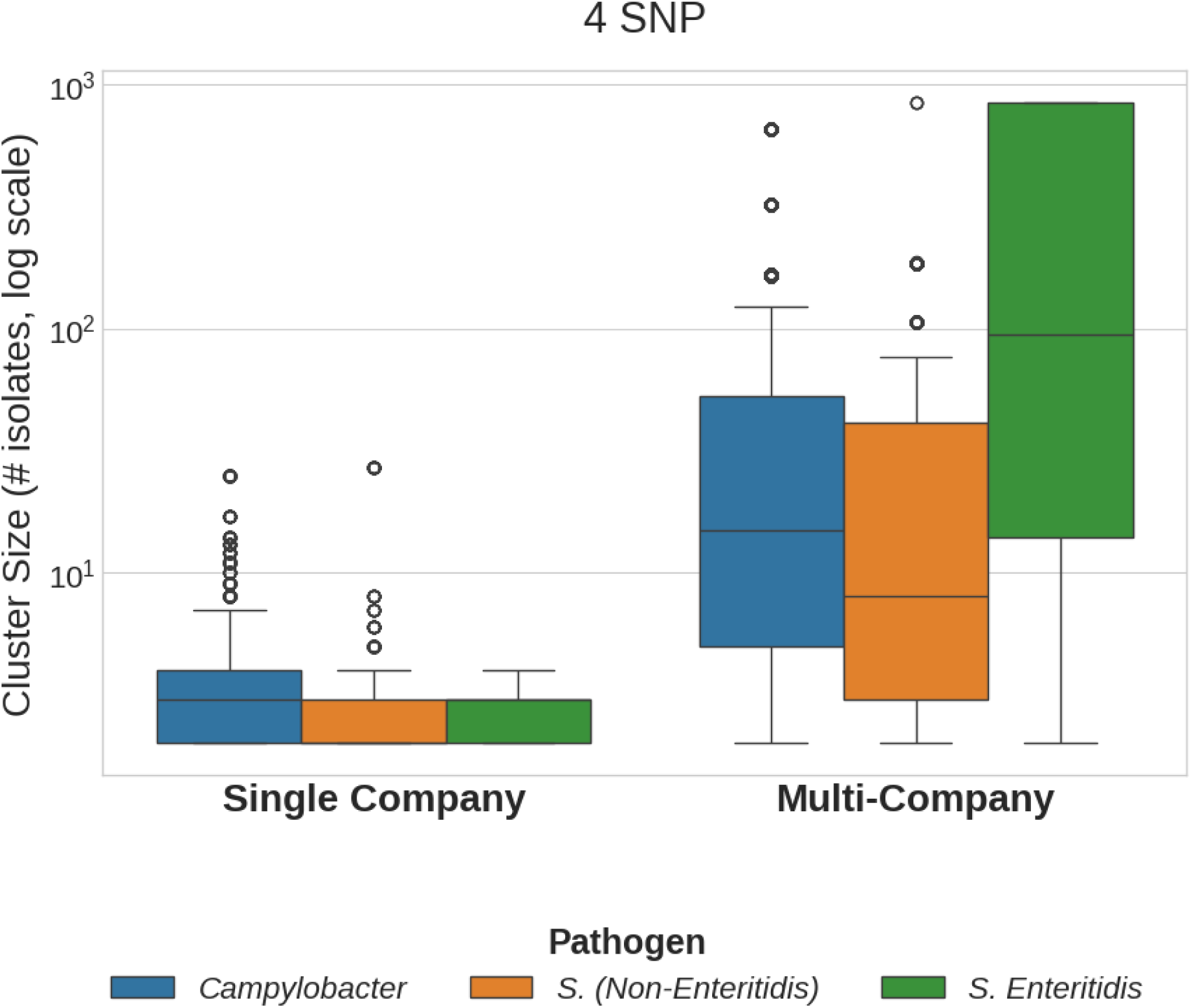
Distribution of Cluster Sizes for Isolates in Single Company versus Multi-Company Clusters. The box plots show the log scale distribution of cluster sizes for isolates in the single company versus multi-company clusters.

For isolates in the same pathogen groups and spread categories as above, **Figure 12** displays boxplots of the distribution of their cluster persistence values, i.e. the time span, in days, between the collection of the cluster’s earliest isolate and its most recent isolate. The multi-company clusters have much longer persistence than single company clusters. Half of the Campylobacter multi-company isolates are in clusters persisting longer than 1250 days, and half of the Enteritidis isolates are in clusters extending throughout the interval of the collection date span. It is notable that even among the single company clusters, a substantial fraction of the isolates are in clusters that persist for at least 250 days which is a longer time period than multiple flock generations in a growout house. Analyses using 2 SNP and 8 SNP thresholds for clustering show qualitatively similar results (**Figure S7**).

**Figure 12.**
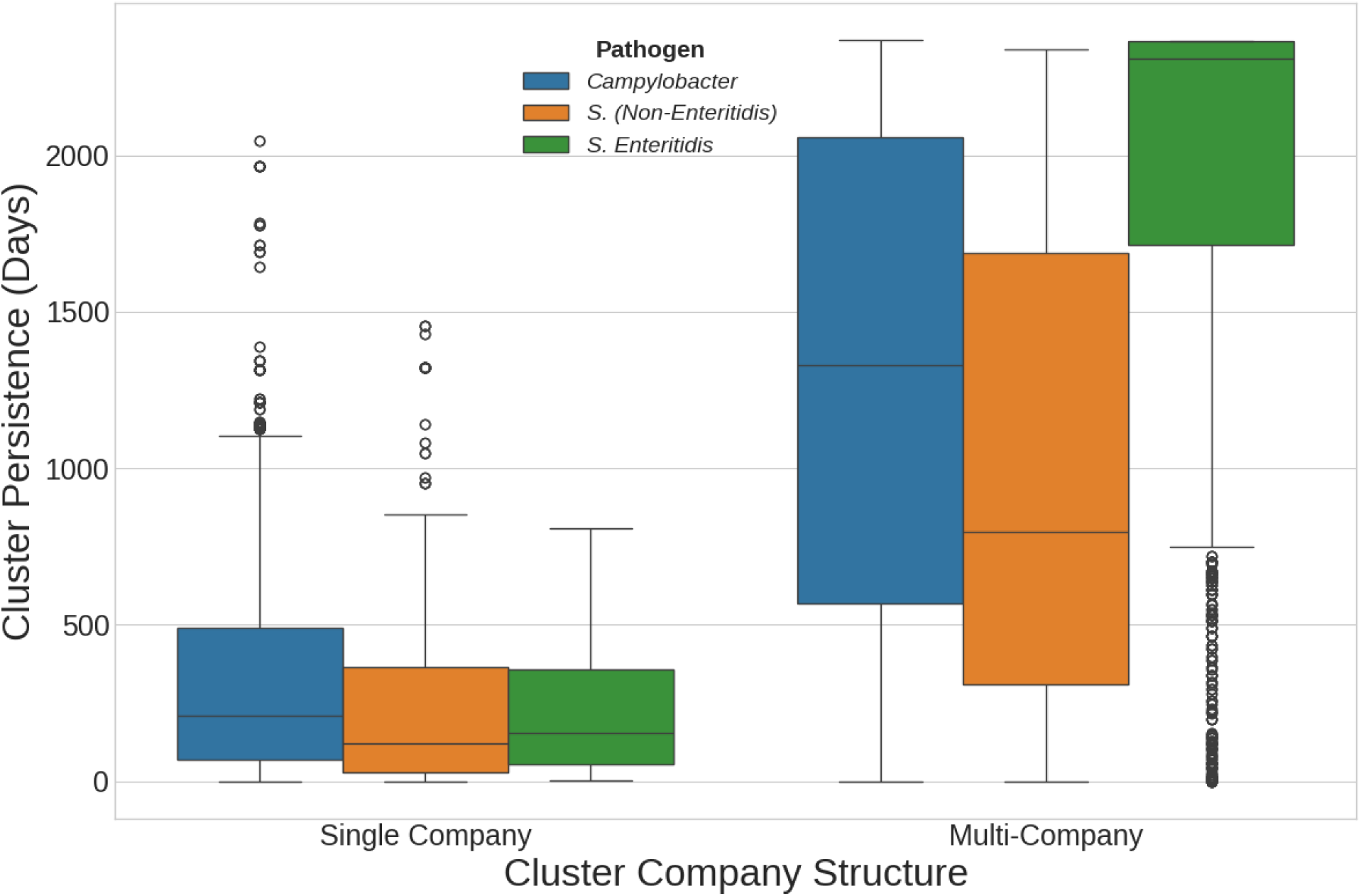
Persistence of Clusters. Distribution of cluster persistence values for isolates in single company versus multi-company clusters, where persistence is defined as the number of days between the collection dates of the first and last samples in the cluster.

Much of the difference in persistence between single and multi-company clusters can be attributed to the much larger cluster sizes for the latter as seen in **Figure 11. Figure S8** compares the persistence of single versus multi-company *Campylobacter* and *Salmonella* clusters for cluster size 2 and cluster size 3. Even with matched cluster sizes, the persistence of multi-company clusters is greater in all cases and statistically significant using the Mann-Whitney U test (Fay and Proschan 2010).

## Discussion

Research into pathogen contamination in broiler production has been extensive, however the FSIS genome sequencing program based on its broiler sampling efforts (“Laboratory Sampling Data,” n.d.) offers a unique opportunity to examine these questions at scale. The combination of longitudinal sampling from over 800 production facilities with the discriminatory capacity of whole-genome sequencing to identify isolates of common origin (Brown et al. 2019; Jagadeesan et al. 2019; Ronholm et al. 2016; E. L. Stevens et al. 2022; Pightling et al. 2018) enables, for the first time, a system-wide assessment of contamination sources in the U.S. broiler production chain.

Analyses of the FSIS sampling data provide compelling evidence that, for the isolates that I can categorize (cluster size > 1), a substantial fraction have their origin at the primary breeder:

1. 83% of Campylobacter isolates, 80% of the *Salmonella* (excluding Enteritidis) isolates, and more than 97% of *Salmonella* Enteritidis isolates are in multi-company clusters (**Figure 8**).
2. For the isolates in multi-company clusters, approximately 93% of *Campylobacter*, 96% of non-Enteritidis *Salmonella*, and 99% of Enteritidis isolates are in geographically dispersed clusters (i.e. maximum or mean pairwise distance > 50 miles). (**Figure 9, Figure S4**).
3. Most of the multi-company isolates are in highly persistent clusters, e.g., for *Campylobacter*, almost 75% of the isolates are in clusters that persist for over 2 years, for non-Enteritidis *Salmonella*, approximately 50% are in clusters that persist for over two years, and more than 75% of the Enteritidis isolates are in clusters that persist for over 4 years.

Given the length of these persistence intervals and the length of our sampling window (less than six years), I am likely to be missing isolates from these clusters that appeared before or after the sampling window. These missing isolates could reveal additional multi-company clusters and thus the results on the proportion of isolates in multi-company clusters, the size of the clusters, and their persistence are conservative; i.e., the missing data can only increase the fraction of contamination likely coming from the primary breeder.

Although the primary breeder is the likely contamination source for most of the isolates of all three pathogen groups, the analyses reveal notable differences among these groups. For Enteritidis, the most epidemiologically significant *Salmonella* serovar in poultry, almost all of the isolates are in multi-company clusters. And its dispersion patterns are nearly identical to the simple random model used for comparison purposes in these analyses (**Figure 10)**. This model assumes there is a single source of contamination with a random distribution of isolates among complexes and companies throughout the United States. It is unclear how one could account for these results with a contamination source downstream of the primary breeder.

Rather, these results are consistent with the contamination originating at the Great GrandParent phase of the primary breeder with subsequent vertical transmission down the breeding pyramid to multiple geographically dispersed GrandParent farms and on to the processing complexes (**Figure 2)**. Furthermore, this close match with the random model indicates that there are minimal differences among companies and processing complexes in eliminating this contamination or in horizontal spread of these strains within complexes. The vertical transmission of Enteritidis in poultry is well established (Keller et al. 1995; De Reu et al. 2006; Berchieri et al. 2001; Bichler, Nagaraja, and Halvorson 1996; Kinde et al. 2000; Louis 1988; R. K. Gast and Beard 1990; Gantois et al. 2009) and the associated systemic infection (Byrd et al. 2025; Richard K. Gast et al. 2022; Sexton et al. 2018) could help explain these results.

As noted above, since there are two primary breeder companies, it is curious that the distribution of the Enteritidis isolates is quite close to that of a random model with a single source. However, given the hierarchical structure of the breeding pyramid (**Figure 1** and **Figure 2)**, it is difficult to deconvolute the signal of the two primary breeder companies without additional data on company-specific supply chains.

Most of the analyzed non-Enteritidis Salmonella and Campylobacter isolates are in multi-company clusters that are geographically dispersed (maximum or mean pairwise distance > 50 miles) and a substantial fraction of these are highly dispersed, 76% and 83% respectively, i.e. clusters with maximum pairwise distances of > 400 miles (**Figure 9, Figure S4)**. However, unlike Enteritidis, 14% of the Campylobacter and 16% of the non-Enteritidis Salmonella isolates are in single complex clusters. Furthermore, as compared with Enteritidis or the random single-source model, the isolates in multi-company clusters show greater regional concentration (**Figure 9, Figure S4,** and **Figure 10).** These results imply that the Great GrandParent stock of the primary breeder is an important source of contamination of these Parent breeders. However, unlike with Enteritidis, these results are consistent with population differences for Campylobacter and non-Enteritidis Salmonella due to differential spread and/or elimination are likely occurring at sites lower in the breeding pyramid, i.e. at the GrandParent farms which serve different geographic regions or at the processing complexes themselves.

Unlike the extensive evidence for direct vertical transmission for Enteritidis in poultry, direct vertical transmission of Infantis, Kentucky, or Typhimurium serovars (the dominant serovars in the non-Enteritidis set) has limited support in literature. However, indirect evidence from the occurrence patterns of similar strains within broiler complexes suggests that Parent breeders are a significant source of contamination for these serovars (Meng et al. 2025; Mughini-Gras et al. 2021; Liljebjelke et al. 2005; Murray et al. 2023).

Conflicting studies exist regarding vertical transmission of *Campylobacter* in poultry (Orhan Sahin, Morishita, and Zhang 2002; N. A. Cox et al. 2012; O. Sahin, Kobalka, and Zhang 2003; Callicott et al. 2006; Pearson et al. 1996; Nelson A. Cox et al. 2002; Newell and Fearnley 2003) and may explain the differences in these results as compared with e.g. Enteritidis.

Nevertheless, regardless of whether transmission is vertical or pseudo-vertical (Cason, Cox, and Bailey 1994), consistent with the results presented here, published studies have documented the presence of identical *Campylobacter* strains across different farms and processing complexes (Frosth et al. 2020; Montero et al. 2024; M. J. A. Stevens et al. 2024; Hiett et al. 2002; Joensen et al. 2025).

A lower bound estimate of the fraction of FSIS verification samples in our analysis (i.e. with cluster size >= 2) with contamination likely originating from the primary breeder is simply the fraction of isolates in multi-company clusters that are geographically dispersed (i.e. with maximum or mean pairwise distance > 50 miles):

- 78% for *Campylobacter* (0.833 * 0.932)
- 77% for non-Enteritidis *Salmonella* (0.803 * 0.964)
- 96% for Enteritidis (0.966 * 0.99)

Given the duration of our sampling window and the persistence of the clusters, I am likely to miss isolates that could reclassify some single company clusters as multi-company clusters. Furthermore, while I cannot categorize isolates in singleton clusters, could some of these isolates also originate from the primary breeder or are these singleton isolates qualitatively distinct from those in larger clusters and only distantly related to the strains found in clusters exceeding size one? Using a threshold of 4 SNPs, 38% of all the *Campylobacter* isolates are singletons but only 24% are more divergent than 8 SNPs from the categorized isolates, and only 12% have more than 25 allelic differences based on whole genome multilocus sequence tagging (wgMLST) used by the NCBI Pathogen Detection resource (“NCBI - Pathogen Detection - NCBI,” n.d.-a). In other words, only a small proportion of *Campylobacter* singletons are significantly divergent from the categorized isolates. For non-Enteritidis *Salmonella,* 70% of the isolates are singletons but only 46% are more divergent than 8 SNPs from the categorized set and only 8% have more than 25 allelic differences from the categorized isolates. And for Enteritidis, 16% are singletons, only 4% of the isolates are more divergent than 8 SNPs, and less than 1% have more than 25 allelic differences from the categorized isolates. It is therefore likely that a substantial percentage of these singletons could also originate from the primary breeder.

In the United States, poultry is the most frequent source of clinical *Salmonella* cases, accounting for 33% according to a recent analysis(Rose et al. 2025). Although the Infantis, Kentucky, and Enteritidis serovars collectively account for over 75% of the *Salmonella* isolates found in our poultry samples, Enteritidis has a dramatically greater impact on human health (**Figure 13**).

**Figure 13.**
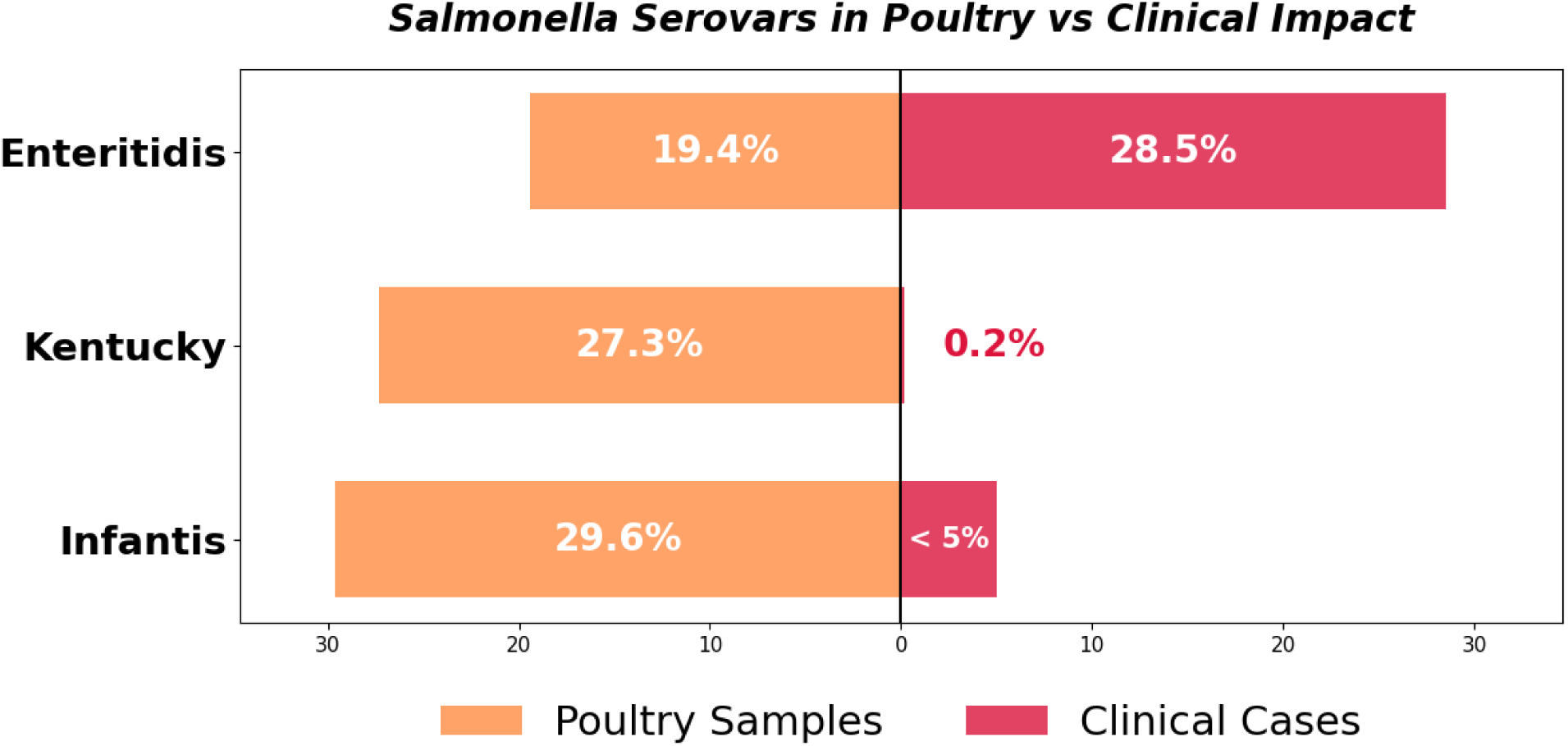
Major *Salmonella* Serovars in Poultry versus Clinical Impact. Orange bars show the relative proportions of Enteritidis, Kentucky, and Infantis serovars among the poultry isolates and the red bars show the relative percentages in US clinical cases (for Enteritidis (Scallan Walter et al. 2025), Infantis (Shah et al. 2024), and Kentucky (Haley et al. 2019)).

Despite ongoing reduction efforts, infection rates from Enteritidis have been relatively stable for at least the last 30 years (“FoodNet” 2023). Our results show that most of the Enteritidis isolates in broilers originate from the earliest stages of the breeding pyramid at the primary breeder, and that differences in the proportion of Enteritidis isolates among processing complexes or companies are insignificant, i.e., mitigation efforts by the broiler processors have had little effect. Given that vertical transmission of Enteritidis is well established in poultry (Keller et al. 1995; De Reu et al. 2006; Berchieri et al. 2001; Bichler, Nagaraja, and Halvorson 1996; Kinde et al. 2000; Louis 1988; R. K. Gast and Beard 1990; Gantois et al. 2009) it is not surprising that, if contamination was introduced early in the breeding pyramid, it would be difficult to reduce or eliminate downstream. It should also be noted however that since a low fraction of the Salmonella and Campylobacter contamination likely originates downstream from the primary breeder, mitigation efforts against contamination originating within the processing complexes themselves have been relatively effective. It is also interesting to note the extremely low fraction of categorizable isolates that are in multi-complex clusters from single companies (**Figure 8).** This implies that efforts by the poultry integrators to reduce cross-contamination between the multiple processing complexes that they own and manage have also been effective.

This study is, to our knowledge, the first to provide quantitative estimates of *Salmonella* and *Campylobacter* contamination percentages originating at the primary breeder stage of broiler production in the United States. Li et al. (2021) proposed that the international dissemination of *Salmonella* Enteritidis was caused by the trade of infected breeding stock, a hypothesis supported by phylogenetic analyses of Enteritidis genomes from various countries (Li et al. 2021). Perhaps the most compelling evidence that the primary breeder is a key contamination source for *Salmonella* comes from the experience of the regulatory agencies in Sweden and Denmark which have virtually eliminated the pathogen from their poultry production systems (Wierup et al. 1995; Wegener et al. 2003; Roberts and Lindblad 2018). The first principle of their control programs involves the quarantining of all imported breeder chicks (e.g. day old GrandParent chicks in the Danish program) to ensure that positives do not enter their production systems. Furthermore, consistent with our results demonstrating the relative importance of vertical or pseudo-vertical versus horizontal transmission in this context, Wegener et al. (2003) states: “In Denmark, most infections appear to be vertically transmitted (nearly always traceable to an infected hatchery or Parent flock), whereas horizontal transmission from the environment and wild fauna appear to play a minor role.” (Wegener et al. 2003).

Controlling *Salmonella* Enteritidis at the primary breeder level, as supported by our findings, promises a substantial reduction in the US illness burden attributed to *Salmonella*. Our data indicate that this pathogen, which is a far greater cause of human illness from broilers than other *Salmonella* serovars(Scallan Walter et al. 2025), originates predominantly in the earliest stages of the primary breeder pyramid and spreads through vertical transmission. Consequently, the number of birds requiring testing and culling at the primary breeder stage is quite small, making this a highly feasible intervention. Furthermore, the effectiveness of this strategy has been demonstrated at a national scale by the successful systems in Sweden and Denmark. Given the significant role of *Salmonella* Enteritidis in overall *Salmonella* illnesses, a measurable improvement in public health is anticipated by targeting this pathogen at its source.

## Materials and Methods

### Datasets

The pathogen isolates used in this project were collected and sequenced by the FSIS as part of their raw poultry verification sampling program and only included the Hazard Analysis and Critical Control Points (HACCP) chicken samples (prefix HC_C for ProjectCode) (“Laboratory Sampling Data,” n.d.; Sampling, n.d.). The sample identifiers and associated metadata used were for samples collected from October 1, 2018 through March 31, 2025. The FSIS IDs were used to link to the associated genome data and SNP distances on the National Center for Biotechnology Information (NCBI) Pathogen Detection site (“NCBI - Pathogen Detection - NCBI,” n.d.-a). The Salmonella serovar classifications used were those assigned by NCBI using SeqSero2 (“NCBI - Pathogen Detection - NCBI,” n.d.-b; Zhang et al. 2019). The latitude and longitude coordinates for the processing complexes were obtained from the FSIS Inspected Establishments directory (“FSIS Inspected Establishments,” n.d.).

### SNP distances, clustering, and thresholds

The SNP distances and clustering were done as described previously (Lipman et al. 2024). A threshold of 4 SNPs was used for the analyses in the Results section and the robustness of these results using thresholds of 2, 4, and 8 SNPs is presented in Supplementary Materials.

### Company Names

In the FSIS JSON metadata, there is an attribute for company name (establishment_name) and in some cases the same company appears with slightly different strings. Using computational tools along with manual review, I have generated a new attribute, “CompanyName” which merges a small number of the EstablishmentName values. The number of unique values in establishment_name is 702, while the number of unique values in CompanyName is 623. The results of our analyses that involve company names include: **Figure 4, Figure 6, Figure 7, Figure 8**, and **Figure 10**. There are only very minimal differences (∼1%) in results using either of these two attributes for company name.

## Supporting information

Salmonella clusters

Campylobacter clusters

## Data Availability

All data is publicly available on the USDA website and at NCBI/NLM/NIH. The links are in the manuscript.

https://www.fsis.usda.gov/news-events/publications/raw-poultry-sampling

https://ftp.ncbi.nlm.nih.gov/pathogen/Results/Salmonella/PDG000000002.3695/

https://ftp.ncbi.nlm.nih.gov/pathogen/Results/Campylobacter/PDG000000003.2583/

## Acknowledgements

I wish to acknowledge the Food Safety Inspection Service - USDA for their efforts in sampling poultry, conducting tests, sequencing, and providing accessible data on *Salmonella* and *Campylobacter* pathogens in poultry. I thank Dr. Steven Musser (Food and Drug Administration), Dr. Errol Strain (Food and Drug Administration), and Dr. Joshua Cherry (National Institutes of Health) for their suggestions and advice. I also thank Dr. Richa Agarwala (National Institutes of Health) for generating the isolate clusters from the original NCBI Pathogen Detection site patristic distances.

## Supplementary Materials

**Figure S1.**
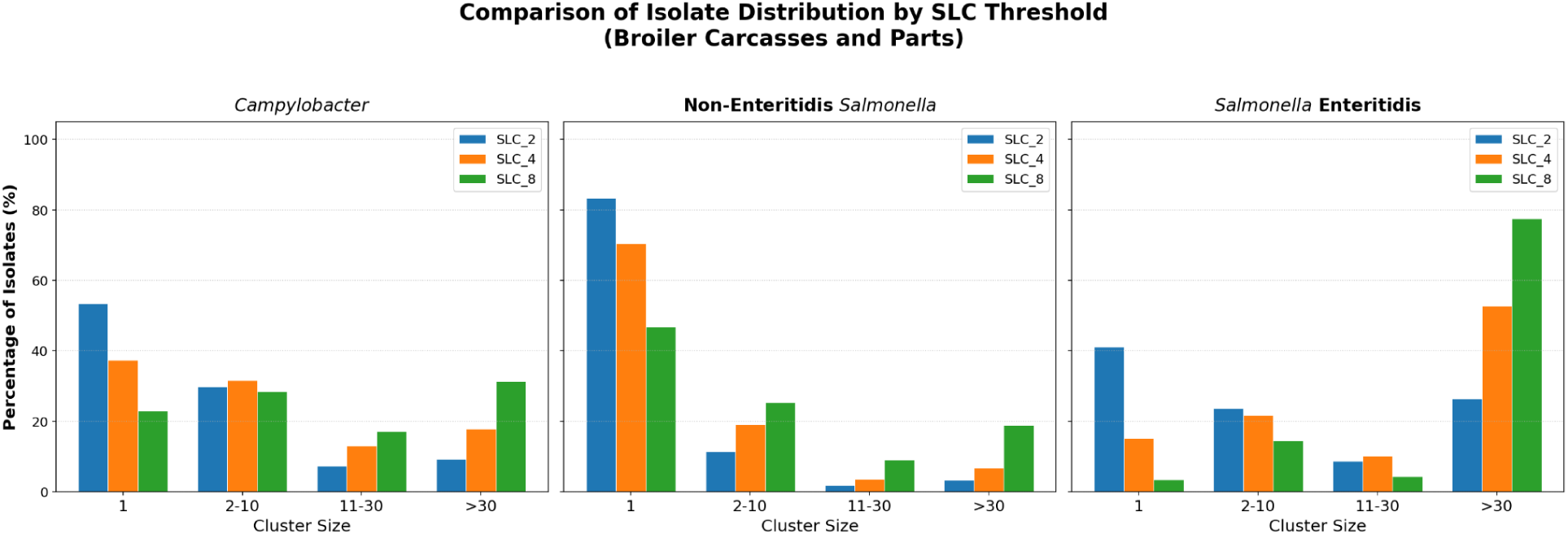
Distribution of isolates by cluster size for *Campylobacter*, non-Enteritidis *Salmonella*, and Enteritidis. Vertical bars represent the proportion of isolates in the cluster size ranges indicated on the x axis for each pathogen group and different SNP thresholds for clustering: 2 SNPs (blue), 4 SNPs (orange), 8 SNPs (green)

**Figure S2.**
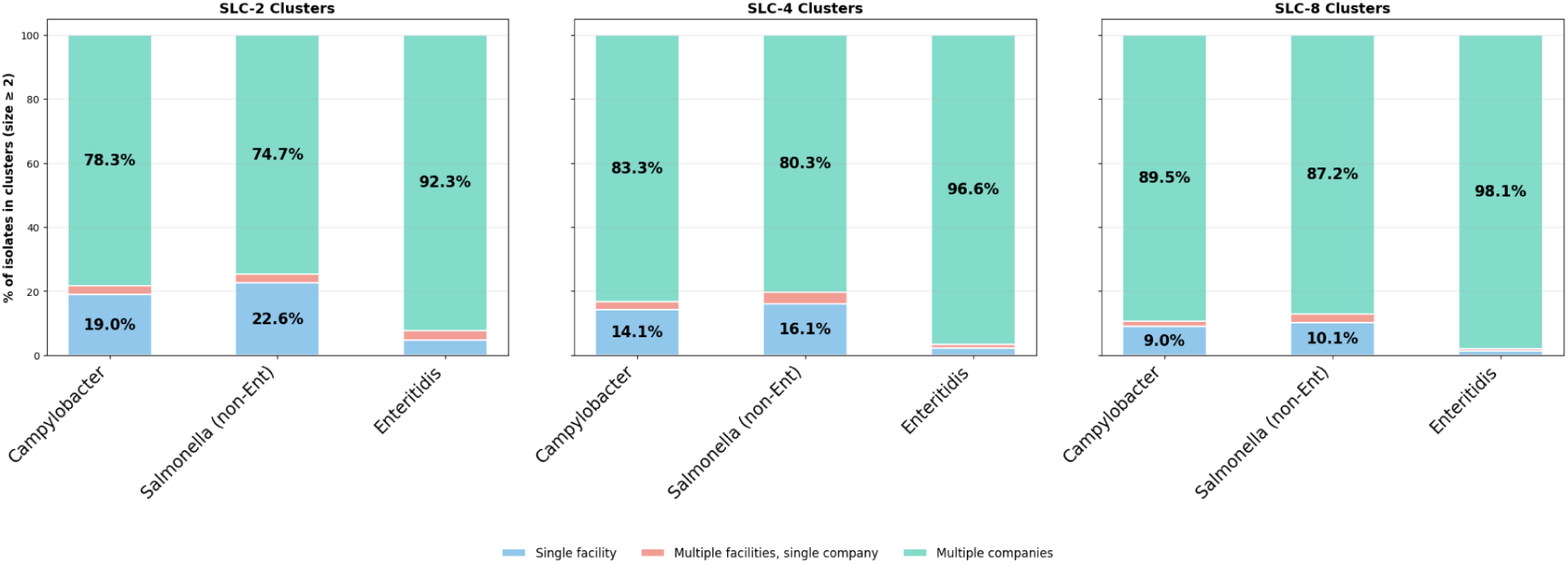
Proportion of isolates in each spread category for *Campylobacter*, Enteritidis, non-Enteritidis *Salmonella*. Stacked bar plots showing the proportion of isolates in single facility, multi-facility and single company, and multi-company clusters for *Campylobacter*, Enteritidis, and non_Enteritidis *Salmonella*. The first panel is for clustering using a 2 SNP threshold, the second panel using a 4 SNP threshold, and the third panel using an 8 SNP threshold.

**Figure S3.**
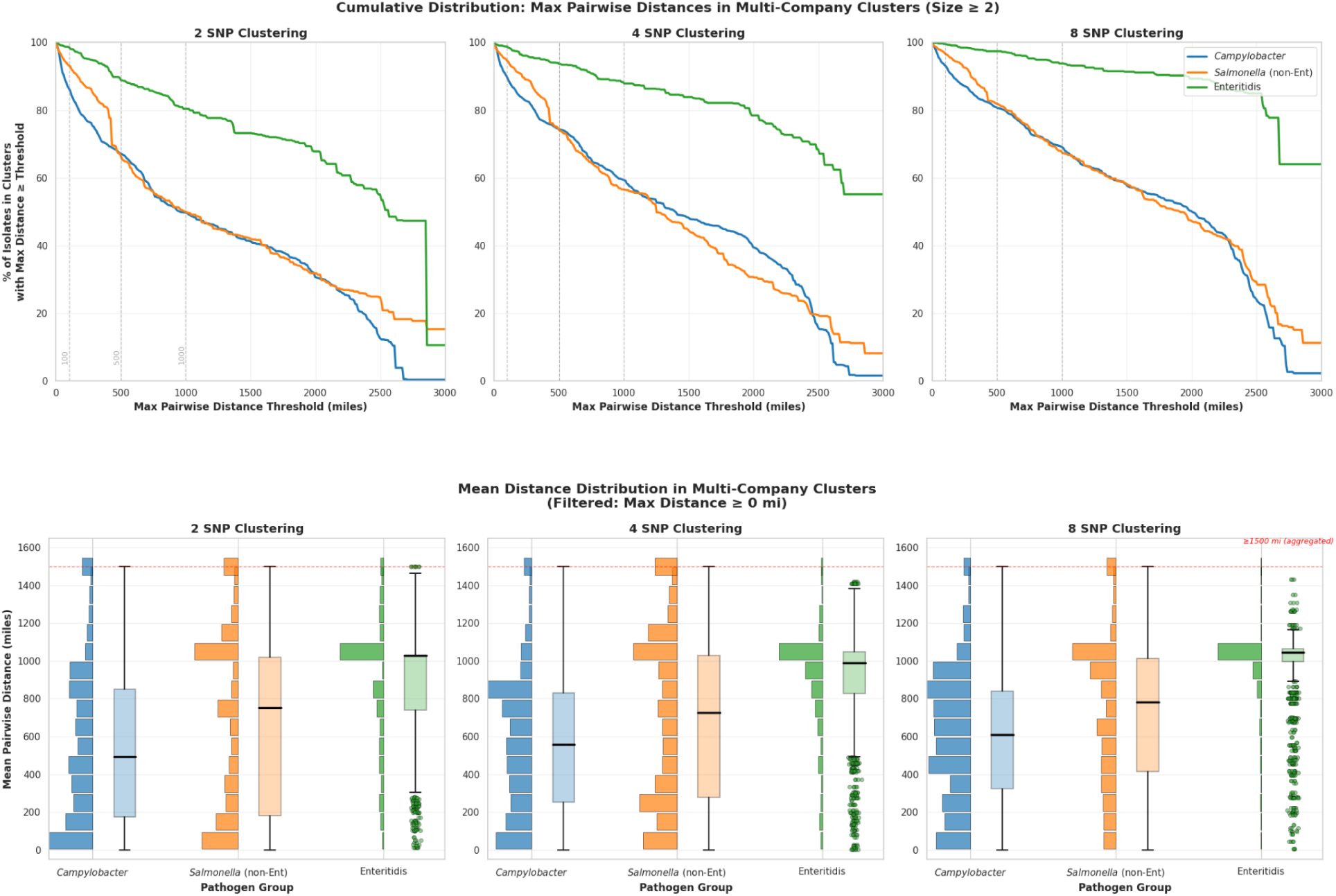
Distribution of Pairwise Distances within Multi-Company Clusters. The upper row shows cumulative distribution curves of the proportion of isolates in clusters with increasing maximum pairwise distance between processing complexes. The three panels show results with 2 SNP, 4 SNP, and 8 SNP clustering thresholds. The bottom row shows histograms and box plots for the distribution of the proportion of isolates in clusters with different mean pairwise distances. The y axis was truncated at 1500 miles and counts for isolates in clusters with mean distance > 1500 are aggregated in the last bin of the histograms. The three panels show results with 2 SNP, 4 SNP, and 8 SNP clustering thresholds.

**Figure S4.**
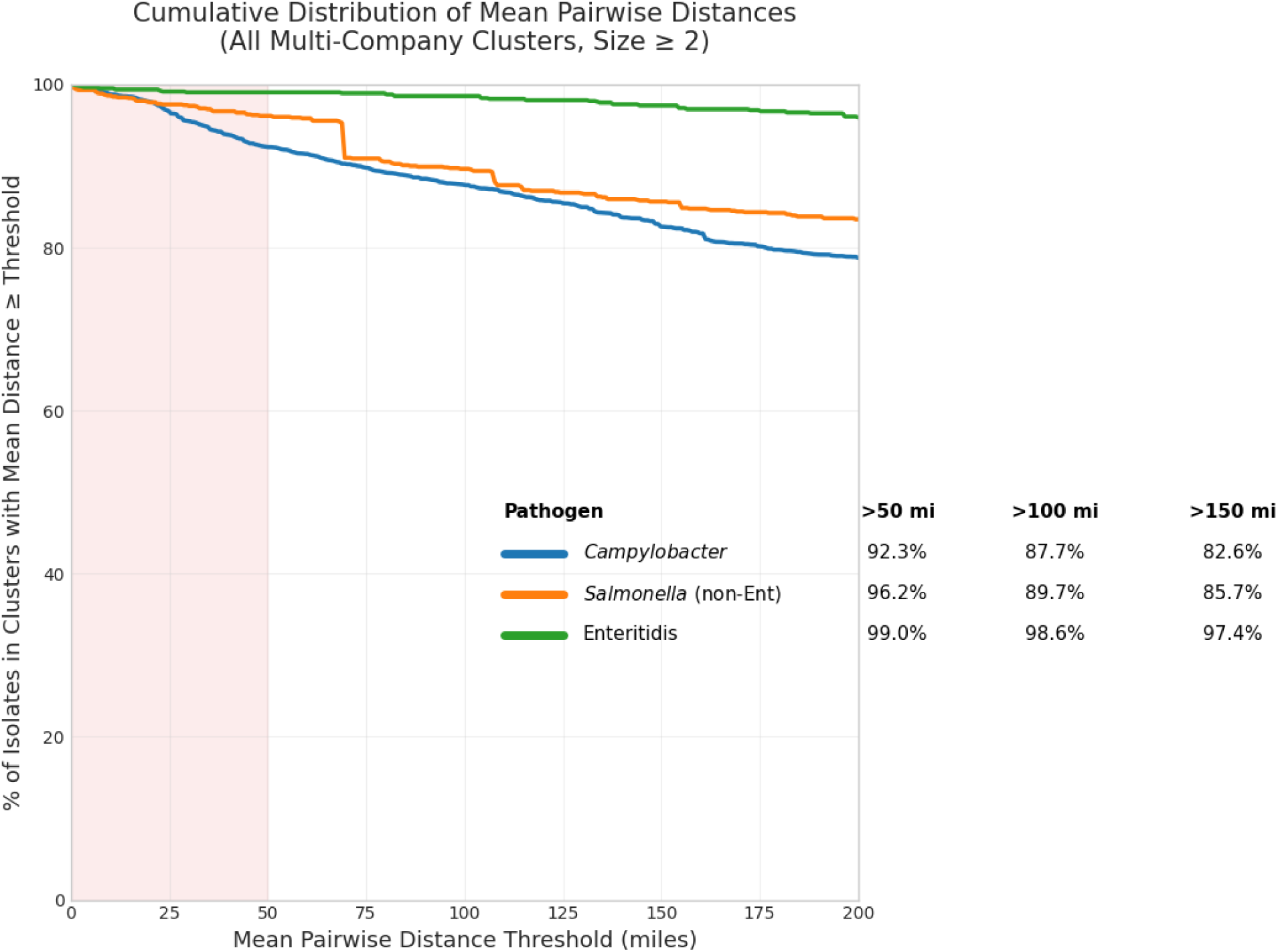
Cumulative Frequency Distribution of Mean Pairwise Distances within Multi-Company Clusters. Cumulative frequency distribution curves of the proportion of isolates in clusters with increasing mean pairwise distance between processing complexes.

**Figure S5.**
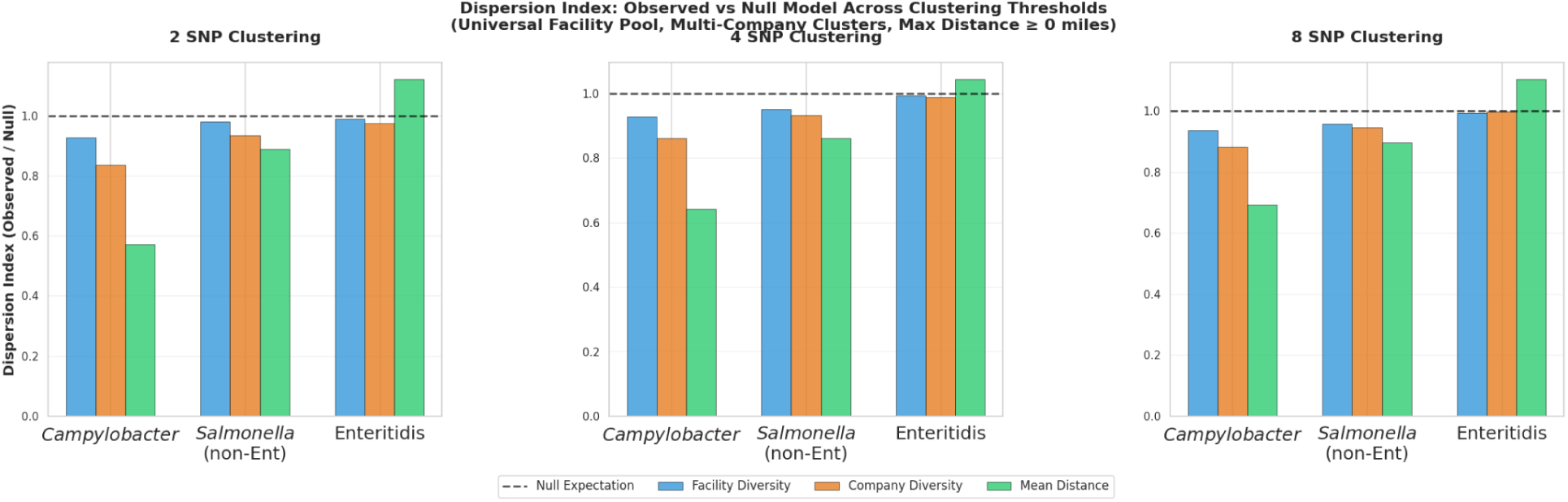
Spread Ratio of Observed versus Random Pair Counts for Multi-Company Clusters. The vertical bars represent the ratios of observed counts versus random pair counts (50iterations): all isolate pairs within each cluster are examined to determine whether they are from different complexes (blue bar) or different companies (orange), and the mean pairwise distance for the cluster is computed. These counts are compared to the same counts for the random reference model. The three panels show the results with a 2 SNP, 4 SNP, and 8 SNP clustering threshold.

**Figure S6.**
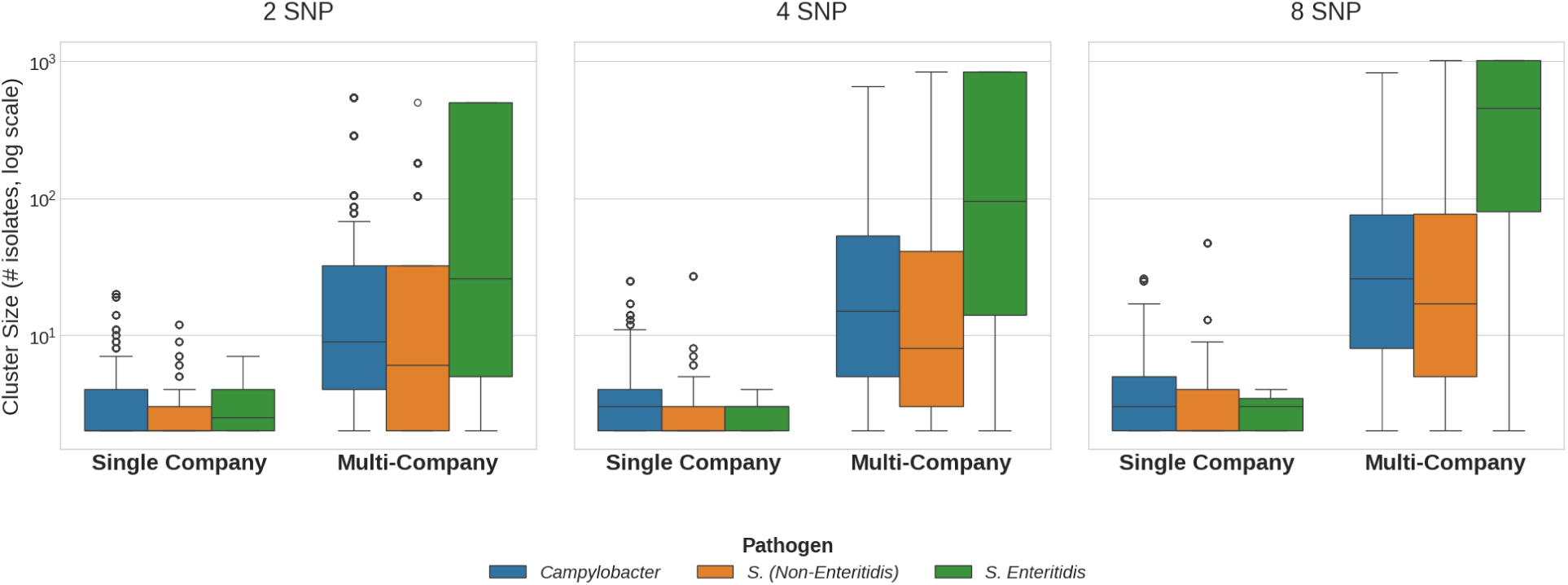
Distribution of Cluster Sizes for Isolates in Single Company versus Multi-Company clusters (2 SNP threshold clustering in left panel, 8 SNP threshold clustering in right panel) The box plots show the log scale distribution of cluster sizes for isolates in the single company versus multi-company clusters.

**Figure S7.**
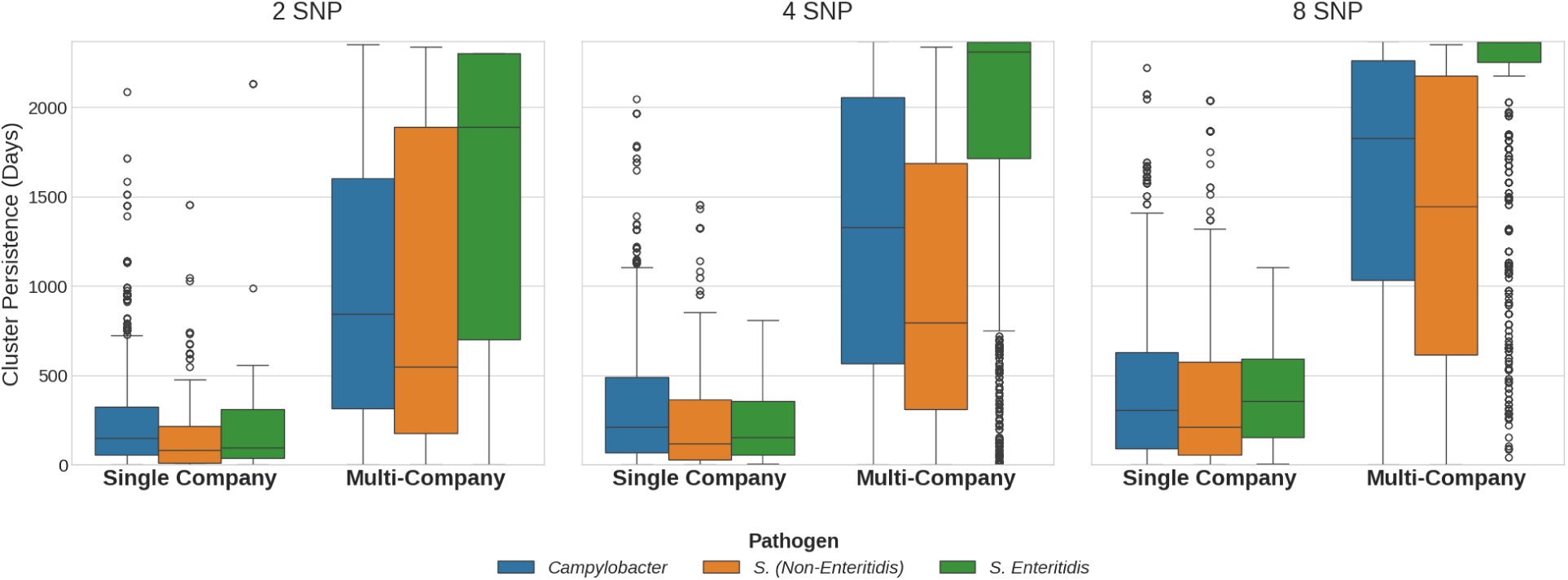
Persistence of Clusters (2 SNP threshold clustering in left panel, 8 SNP threshold clustering in right panel) Distribution of cluster persistence values for isolates in single company versus multi-company clusters, where persistence is defined as the number of days between the collection dates of the first and last samples in the cluster.

**Figure S8.**
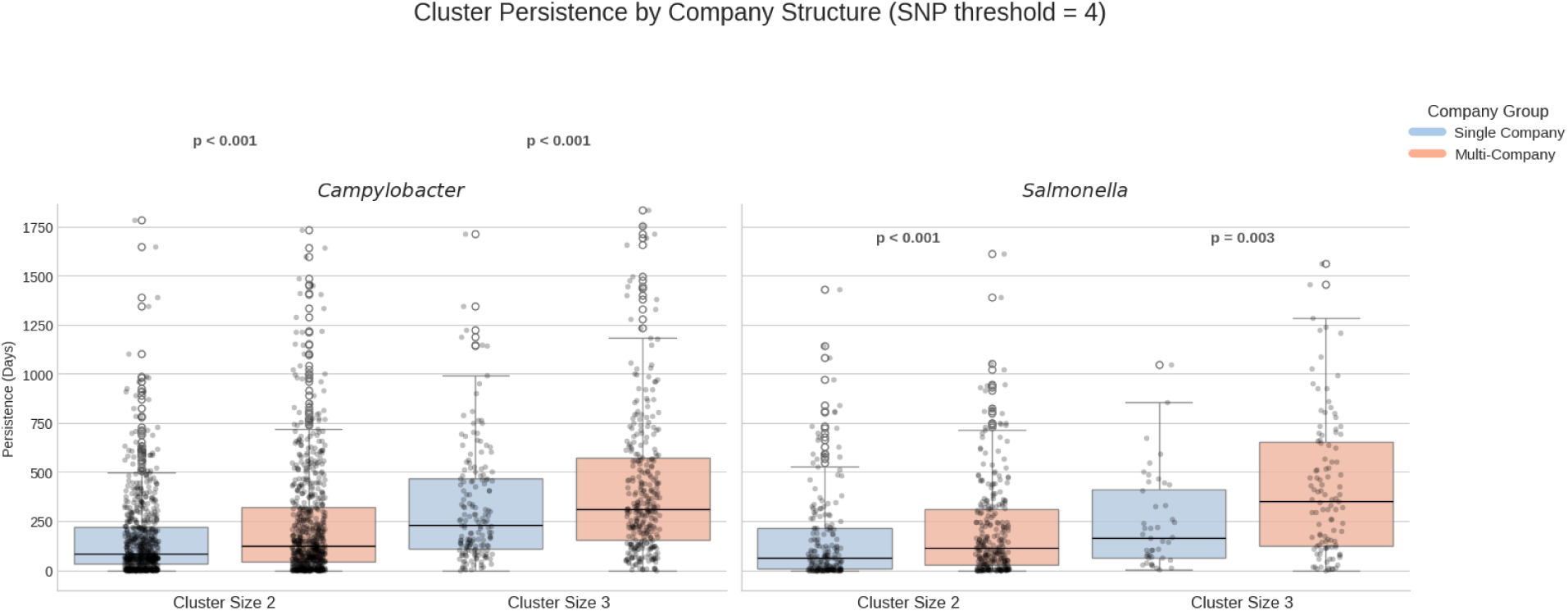
Comparison of Persistence for Single Company vs Multi-Company Clusters for Clusters of Size 2 and Size 3. Box plots with overlaid strip plots for persistence of clusters of size 2 and size 3 with Single Company (blue) vs Multi-Company clusters (orange). P values are computed using Mann-Whitney U test.

## References

Akilian, Harout. 2020. “New Player Eyeing to Break in the U.S Broiler Duopoly with Improved Breed.” aviNews. agriNews. August 17, 2020. https://avinews.com/en/new-player-eyeing-to-break-in-the-us-broiler-duopoly-with-improved-breed/.

“Animal Health.” n.d. Accessed July 4, 2025. https://www.poultryimprovement.org/.

Berchieri, A., Jr, P. Wigley, K. Page, C. K. Murphy, and P. A. Barrow. 2001. “Further Studies on Vertical Transmission and Persistence of Salmonella Enterica Serovar Enteritidis Phage Type 4 in Chickens.” Avian Pathology: Journal of the W.V.P.A 30 (4): 297–310.

Bichler, L. A., K. V. Nagaraja, and D. A. Halvorson. 1996. “Salmonella Enteritidis in Eggs, Cloacal Swab Specimens, and Internal Organs of Experimentally Infected White Leghorn Chickens.” American Journal of Veterinary Research 57 (4): 489–95.

Brown, Eric, Uday Dessai, Sherri McGarry, and Peter Gerner-Smidt. 2019. “Use of Whole-Genome Sequencing for Food Safety and Public Health in the United States.” Foodborne Pathogens and Disease 16 (7): 441–50.

Byrd, J. Allen, Sarah Faust, Denise Y. Caldwell, Christina L. Swaggerty, Kenneth Genovese, Micheal H. Kogut, Anna V. Carlson, Casey Johnson, Toni Poole, and Keri N. Norman. 2025. “Recovery of Salmonella from Alternative Anatomical Sites after an Oral Challenge with Three Different Salmonella Serotypes in Turkeys.” Poultry Science 104 (9): 105406.

Callicott, Kenneth A., Vala Friethriksdóttir, Jarle Reiersen, Ruff Lowman, Jean-Robert Bisaillon, Eggert Gunnarsson, Eva Berndtson, Kelli L. Hiett, David S. Needleman, and Norman J. Stern. 2006. “Lack of Evidence for Vertical Transmission of Campylobacter Spp. in Chickens.” Applied and Environmental Microbiology 72 (9): 5794–98.

Cason, J. A., N. A. Cox, and J. S. Bailey. 1994. “Transmission of Salmonella Typhimurium during Hatching of Broiler Chicks.” Avian Diseases 38 (3): 583–88.

CDC. 2025. “Foodborne Illness Source Attribution Estimates – United States, 2022.” Interagency Food Safety Analytics Collaboration. January 16, 2025. https://www.cdc.gov/ifsac/php/data-research/annual-report-2022.html.

Chavez-Velado, Daniela R., David A. Vargas, and Marcos X. Sanchez-Plata. 2024. “Bio-Mapping Salmonella and Campylobacter Loads in Three Commercial Broiler Processing Facilities in the United States to Identify Strategic Intervention Points.” Foods (Basel, Switzerland) 13 (2): 180.

Cody, Alison J., Martin Cj Maiden, Norval Jc Strachan, and Noel D. McCarthy. 2019. “A Systematic Review of Source Attribution of Human Campylobacteriosis Using Multilocus Sequence Typing.” Euro Surveillance : Bulletin Europeen Sur Les Maladies Transmissibles [Euro Surveillance : European Communicable Disease Bulletin] 24 (43): 1800696.

Cox, N. A., L. J. Richardson, J. J. Maurer, M. E. Berrang, P. J. Fedorka-Cray, R. J. Buhr, J. A. Byrd, et al. 2012. “Evidence for Horizontal and Vertical Transmission in Campylobacter Passage from Hen to Her Progeny.” Journal of Food Protection 75 (10): 1896–1902.

Cox, Nelson A., Norman J. Stern, Kelli L. Hiett, and Mark E. Berrang. 2002. “Identification of a New Source of Campylobacter Contamination in Poultry: Transmission from Breeder Hens to Broiler Chickens.” Avian Diseases 46 (3): 535–41.

De Reu, K., K. Grijspeerdt, W. Messens, M. Heyndrickx, M. Uyttendaele, J. Debevere, and L. Herman. 2006. “Eggshell Factors Influencing Eggshell Penetration and Whole Egg Contamination by Different Bacteria, Including Salmonella Enteritidis.” International Journal of Food Microbiology 112 (3): 253–60.

Fay, Michael P., and Michael A. Proschan. 2010. “Wilcoxon-Mann-Whitney or T-Test? On Assumptions for Hypothesis Tests and Multiple Interpretations of Decision Rules.” Statistics Surveys 4:1–39.

“Federal Meat Inspection Act.” n.d. Accessed April 10, 2025. https://www.fsis.usda.gov/policy/food-safety-acts/federal-meat-inspection-act.

“FoodNet.” 2023. June 28, 2023. https://www.cdc.gov/foodnet/index.html.

Frosth, Sara, Oskar Karlsson-Lindsjö, Adnan Niazi, Lise-Lotte Fernström, and Ingrid Hansson. 2020. “Identification of Transmission Routes of Campylobacter and on-Farm Measures to Reduce Campylobacter in Chicken.” Pathogens 9 (5): 363.

“FSIS Compliance Guideline: Modernization of Poultry Slaughter Inspection Microbiological Sampling of Raw Poultry.” 2015. https://www.fsis.usda.gov/sites/default/files/import/Microbiological-Testing-Raw-Poultry.pdf.

“FSIS Inspected Establishments.” n.d. Accessed November 3, 2025. https://www.fsis.usda.gov/inspection/fsis-inspected-establishments.

Gantois, Inne, Richard Ducatelle, Frank Pasmans, Freddy Haesebrouck, Richard Gast, Tom J. Humphrey, and Filip Van Immerseel. 2009. “Mechanisms of Egg Contamination by Salmonella Enteritidis.” FEMS Microbiology Reviews 33 (4): 718–38.

Gast, Richard K., Deana R. Jones, Rupa Guraya, Javier S. Garcia, and Darrin M. Karcher. 2022. “Research Note: Internal Organ Colonization by Salmonella Enteritidis in Experimentally Infected Layer Pullets Reared at Different Stocking Densities in Indoor Cage-Free Housing.” Poultry Science 101 (11): 102104.

Gast, R. K., and C. W. Beard. 1990. “Production of Salmonella Enteritidis-Contaminated Eggs by Experimentally Infected Hens.” Avian Diseases 34 (2): 438–46.

Golden, Chase E., and Abhinav Mishra. 2020. “Prevalence of Salmonella and Campylobacter Spp. In Alternative and Conventionally Produced Chicken in the United States: A Systematic Review and Meta-Analysis.” Journal of Food Protection 83 (7): 1181–97.

Haley, Bradd J., Seon Woo Kim, Julie Haendiges, Eric Keller, David Torpey, Alexander Kim, Kia Crocker, Robert A. Myers, and Jo Ann S. Van Kessel. 2019. “Salmonella Enterica Serovar Kentucky Recovered from Human Clinical Cases in Maryland, USA (2011-2015).” Zoonoses and Public Health 66 (4): 382–92.

Hiett, K. L., N. J. Stern, P. Fedorka-Cray, N. A. Cox, M. T. Musgrove, and S. Ladely. 2002. “Molecular Subtype Analyses of Campylobacter Spp. from Arkansas and California Poultry Operations.” Applied and Environmental Microbiology 68 (12): 6220–36.

Iowa State University, {Animal and Plant Health Inspection Service], and USDA. n.d. “US Poultry Industry Manual.” Accessed April 10, 2025. https://www.aphis.usda.gov/sites/default/files/poultry_ind_manual.pdf.

Jackson, Brendan R., Cheryl Tarr, Errol Strain, Kelly A. Jackson, Amanda Conrad, Heather Carleton, Lee S. Katz, et al. 2016. “Implementation of Nationwide Real-Time Whole-Genome Sequencing to Enhance Listeriosis Outbreak Detection and Investigation.” Clinical Infectious Diseases: An Official Publication of the Infectious Diseases Society of America 63 (3): 380–86.

Jagadeesan, Balamurugan, Peter Gerner-Smidt, Marc W. Allard, Sébastien Leuillet, Anett Winkler, Yinghua Xiao, Samuel Chaffron, et al. 2019. “The Use of next Generation Sequencing for Improving Food Safety: Translation into Practice.” Food Microbiology 79 (June):96–115.

Joensen, Katrine Grimstrup, Gitte Sørensen, Pernille Gymoese, Louise Gade Dahl, and Eva Møller Nielsen. 2025. “The Clonal Spread and Persistence of Campylobacter in Danish Broiler Farms and Its Association with Human Infections.” Pathogens 14 (8): 821.

Johnston, J. 2020. “Food Safety and Inspection Service.” Federal Regulatory Guide. 10.4135/9781544377230.n47.

Keller, L. H., C. E. Benson, K. Krotec, and R. J. Eckroade. 1995. “Salmonella Enteritidis Colonization of the Reproductive Tract and Forming and Freshly Laid Eggs of Chickens.” Infection and Immunity 63 (7): 2443–49.

Kinde, H., H. L. Shivaprasad, B. M. Daft, D. H. Read, A. Ardans, R. Breitmeyer, G. Rajashekara, K. V. Nagaraja, and I. A. Gardner. 2000. “Pathologic and Bacteriologic Findings in 27-Week-Old Commercial Laying Hens Experimentally Infected with Salmonella Enteritidis, Phage Type 4.” Avian Diseases 44 (2): 239–48.

“Laboratory Sampling Data.” n.d. Accessed April 10, 2025. https://www.fsis.usda.gov/science-data/data-sets-visualizations/laboratory-sampling-data.

Liljebjelke, Karen A., Charles L. Hofacre, Tongrui Liu, David G. White, Sherry Ayers, Suzanne Young, and John J. Maurer. 2005. “Vertical and Horizontal Transmission of Salmonella within Integrated Broiler Production System.” Foodborne Pathogens and Disease 2 (1): 90–102.

Lipman, David J., Joshua L. Cherry, Errol Strain, Richa Agarwala, and Steven M. Musser. 2024. “Genomic Perspectives on Foodborne Illness.” Proceedings of the National Academy of Sciences of the United States of America 121 (46): e2411894121.

Li, Shaoting, Yingshu He, David Ames Mann, and Xiangyu Deng. 2021. “Global Spread of Salmonella Enteritidis via Centralized Sourcing and International Trade of Poultry Breeding Stocks.” Nature Communications 12 (1): 5109.

Louis, Michael E. St. 1988. “The Emergence of Grade A Eggs as a Major Source of Salmonella Enteritidis Infections: New Implications for the Control of Salmonellosis.” JAMA: The Journal of the American Medical Association 259 (14): 2103.

MacDonald, James M. 2008. “The Economic Organization of U.S. Broiler Production.” https://ers.usda.gov/sites/default/files/_laserfiche/publications/44254/12067_eib38_1_.pdf.

Meng, Chuang, Fan Wang, Chen Xu, Bowen Liu, Xilong Kang, Yunzeng Zhang, Xinan Jiao, and Zhiming Pan. 2025. “Prevalence and Transmission of Salmonella Collected from Farming to Egg Processing of Layer Production Chain in Jiangsu Province, China.” Poultry Science 104 (2): 104714.

Montero, Lorena, José L. Medina-Santana, María Ishida, Brian Sauders, Gabriel Trueba, and Christian Vinueza-Burgos. 2024. “Transmission of Dominant Strains of Campylobacter Jejuni and Campylobacter Coli between Farms and Retail Stores in Ecuador: Genetic Diversity and Antimicrobial Resistance.” PloS One 19 (9): e0308030.

Mughini-Gras, Lapo, Angela H. A. M. van Hoek, Tryntsje Cuperus, Cecile Dam-Deisz, Wendy van Overbeek, Maaike van den Beld, Ben Wit, et al. 2021. “Prevalence, Risk Factors and Genetic Traits of Salmonella Infantis in Dutch Broiler Flocks.” Veterinary Microbiology 258 (109120): 109120.

Murray, Carolyn E., Csaba Varga, Rachel Ouckama, and Michele T. Guerin. 2023. “Temporal Study of Salmonella Enterica Serovars Isolated from Environmental Samples from Ontario Poultry Breeder Flocks between 2009 and 2018.” Pathogens 12 (2): 278.

National Poultry Improvement Plan. n.d. “NPIP Program Standards.” https://poultryimprovement.org/documents/standarde-biosecurityprinciples.pdf.

“NCBI - Pathogen Detection - NCBI.” n.d.-a. Accessed August 25, 2025. https://www.ncbi.nlm.nih.gov/pathogens/docs/data_processing/#data-processing-clustering.

“NCBI - Pathogen Detection - NCBI.” n.d.-b. Accessed September 17, 2025. https://www.ncbi.nlm.nih.gov/pathogens/pathogens_help/#seqsero2_2019.

Newell, D. G., and C. Fearnley. 2003. “Sources of Campylobacter Colonization in Broiler Chickens.” Applied and Environmental Microbiology 69 (8): 4343–51.

Pascoe, Ben, Georgina Futcher, Johan Pensar, Sion C. Bayliss, Evangelos Mourkas, Jessica K. Calland, Matthew D. Hitchings, et al. 2024. “Machine Learning to Attribute the Source of Campylobacter Infections in the United States: A Retrospective Analysis of National Surveillance Data.” The Journal of Infection 89 (5): 106265.

Pearson, A. D., M. H. Greenwood, R. K. Feltham, T. D. Healing, J. Donaldson, D. M. Jones, and R. R. Colwell. 1996. “Microbial Ecology of Campylobacter Jejuni in a United Kingdom Chicken Supply Chain: Intermittent Common Source, Vertical Transmission, and Amplification by Flock Propagation.” Applied and Environmental Microbiology 62 (12): 4614–20.

Pightling, Arthur W., James B. Pettengill, Yan Luo, Joseph D. Baugher, Hugh Rand, and Errol Strain. 2018. “Interpreting Whole-Genome Sequence Analyses of Foodborne Bacteria for Regulatory Applications and Outbreak Investigations.” Frontiers in Microbiology 9 (July):1482.

Pollock, D. L. 1999. “A Geneticist’s Perspective from within a Broiler Primary Breeder Company.” Poultry Science 78 (3): 414–18.

Roberts, Tanya, and Johan Lindblad. 2018. “Sweden Led Salmonella Control in Broilers: Which Countries Are Following?” In Food Safety Economics, 231–49. Cham: Springer International Publishing.

Ronholm, J., Neda Nasheri, Nicholas Petronella, and Franco Pagotto. 2016. “Navigating Microbiological Food Safety in the Era of Whole-Genome Sequencing.” Clinical Microbiology Reviews 29 (4): 837–57.

Rose, Erica Billig, Molly K. Steele, Beth Tolar, James Pettengill, Michael Batz, Michael Bazaco, Berhanu Tameru, et al. 2025. “Attribution of Salmonella Enterica to Food Sources by Using Whole-Genome Sequencing Data.” Emerging Infectious Diseases 31 (4): 783–90.

Sahin, O., P. Kobalka, and Q. Zhang. 2003. “Detection and Survival of Campylobacter in Chicken Eggs.” Journal of Applied Microbiology 95 (5): 1070–79.

Sahin, Orhan, Teresa Y. Morishita, and Qijing Zhang. 2002. “Campylobacter Colonization in Poultry: Sources of Infection and Modes of Transmission.” Animal Health Research Reviews 3 (2): 95–105.

Sampling, Raw Poultry. n.d. “Raw Poultry Sampling.” Accessed September 4, 2025. https://www.fsis.usda.gov/news-events/publications/raw-poultry-sampling.

Scallan Walter, Elaine J., Zhaohui Cui, Reese Tierney, Patricia M. Griffin, Robert M. Hoekstra, Daniel C. Payne, Erica B. Rose, et al. 2025. “Foodborne Illness Acquired in the United States-Major Pathogens, 2019.” Emerging Infectious Diseases 31 (4): 669–77.

Sexton, T. Y., Ifigenia Geornaras, Keith E. Belk, Marisa Bunning, and Jennifer N. Martin. 2018. “Salmonella Contamination in Broiler Synovial Fluid: Are We Missing a Potential Reservoir?” Journal of Food Protection 81 (9): 1425–31.

Shah, Hazel J., Rachel H. Jervis, Katie Wymore, Tamara Rissman, Bethany LaClair, Michelle M. Boyle, Kirk Smith, et al. 2024. “Reported Incidence of Infections Caused by Pathogens Transmitted Commonly Through Food: Impact of Increased Use of Culture-Independent Diagnostic Tests - Foodborne Diseases Active Surveillance Network, 1996-2023.” MMWR. Morbidity and Mortality Weekly Report 73 (26): 584–93.

Stevens, Eric L., Heather A. Carleton, Jennifer Beal, Glenn E. Tillman, Rebecca L. Lindsey, A. C. Lauer, Arthur Pightling, et al. 2022. “Use of Whole Genome Sequencing by the Federal Interagency Collaboration for Genomics for Food and Feed Safety in the United States.” Journal of Food Protection 85 (5): 755–72.

Stevens, Marc J. A., Roger Stephan, Jule Anna Horlbog, Nicole Cernela, and Magdalena Nüesch-Inderbinen. 2024. “Whole Genome Sequence-Based Characterization of Campylobacter Isolated from Broiler Carcasses over a Three-Year Period in a Big Poultry Slaughterhouse Reveals High Genetic Diversity and a Recurring Genomic Lineage of Campylobacter Jejuni.” Infection, Genetics and Evolution: Journal of Molecular Epidemiology and Evolutionary Genetics in Infectious Diseases 119 (105578): 105578.

United States Department of Agriculture. n.d. “Poultry Slaughter 2024 Summary.” https://downloads.usda.library.cornell.edu/usda-esmis/files/pg15bd88s/k356c167m/z029r057b/pslaan25.pdf.

“U.S.C. Title 21 - FOOD AND DRUGS.” n.d. Accessed April 10, 2025. https://www.govinfo.gov/content/pkg/USCODE-2021-title21/html/USCODE-2021-title21-chap12-subchapI-sec604.htm.

Usda, Aphis. n.d. “Checklist for Cleaning and Disinfecting Poultry Enclosures.” https://www.aphis.usda.gov/sites/default/files/fsc-birds-checklist-english.pdf.

USDA/FSIS. n.d. “Food Safety and Inspection Service Annual Sampling Plan Fiscal Year 2024.” https://www.fsis.usda.gov/sites/default/files/media_file/documents/FSIS-Annual-Sampling-Plan-FY2024.pdf.

USDA, Iowa State University Center for Food Security and Public Health. 2013. “POULTRY INDUSTRY MANUAL, USDA.” https://www.aphis.usda.gov/sites/default/files/poultry_ind_manual.pdf.

USDA, The Center for Food Security and Public Health, Iowa State University. n.d. “Information Manual for Implementing Poultry Biosecurity.” https://poultrybiosecurity.org/files/Poultry-Biosecurity-Info-Manual.pdf.

Van Eenennaam, Alison L., Kent A. Weigel, Amy E. Young, Matthew A. Cleveland, and Jack C. M. Dekkers. 2014. “Applied Animal Genomics: Results from the Field.” Annual Review of Animal Biosciences 2 (1): 105–39.

Wang, J., S. Vaddu, S. Bhumanapalli, A. Mishra, T. Applegate, M. Singh, and H. Thippareddi. 2023a. “A Systematic Review and Meta-Analysis of the Sources of Campylobacter in Poultry Production (preharvest) and Their Relative Contributions to the Microbial Risk of Poultry Meat.” Poultry Science 102 (10): 102905.

Wang, J., S. Vaddu, S. Bhumanapalli, A. Mishra, T. Applegate, M. Singh, and H. Thippareddi. 2023b. “A Systematic Review and Meta-Analysis of the Sources of Salmonella in Poultry Production (pre-Harvest) and Their Relative Contributions to the Microbial Risk of Poultry Meat.” Poultry Science 102 (5): 102566.

Wegener, Henrik C., Tine Hald, Danilo Lo Fo Wong, Mogens Madsen, Helle Korsgaard, Flemming Bager, Peter Gerner-Smidt, and Kåre Mølbak. 2003. “Salmonella Control Programs in Denmark.” Emerging Infectious Diseases 9 (7): 774–80.

Wierup, M., B. Engström, A. Engvall, and H. Wahlström. 1995. “Control of Salmonella Enteritidis in Sweden.” International Journal of Food Microbiology 25 (3): 219–26.

Williams, Michael S., Eric D. Ebel, Kis Robertson-Hale, Sheryl L. Shaw, and Bonnie W. Kissler. 2025. “Differences in Salmonella Serotypes in Broiler Chickens within and between Slaughter Establishments in the United States.” Journal of Food Protection 88 (6): 100506.

Wilson, Daniel J., Edith Gabriel, Andrew J. H. Leatherbarrow, John Cheesbrough, Steven Gee, Eric Bolton, Andrew Fox, Paul Fearnhead, C. Anthony Hart, and Peter J. Diggle. 2008. “Tracing the Source of Campylobacteriosis.” PLoS Genetics 4 (9): e1000203.

Zhang, Shaokang, Hendrik C. den Bakker, Shaoting Li, Jessica Chen, Blake A. Dinsmore, Charlotte Lane, A. C. Lauer, Patricia I. Fields, and Xiangyu Deng. 2019. “SeqSero2: Rapid and Improved Salmonella Serotype Determination Using Whole-Genome Sequencing Data.” Applied and Environmental Microbiology 85 (23): e01746–19.

